# Land Use Change and Coronavirus Emergence Risk

**DOI:** 10.1101/2020.07.31.20166090

**Authors:** Maria Cristina Rulli, Paolo D’Odorico, Nikolas Galli, David T.S. Hayman

## Abstract

Coronavirus disease 2019 (COVID-19) and severe acute respiratory syndrome (SARS) causing coronaviruses are mostly discovered in Asian horseshoe bats. It is still unclear how ongoing land use changes may facilitate SARS-related coronavirus transmission to humans. Here we use a multivariate hotspot analysis of high-resolution land-use data to show that regions of China populated by horseshoe bats are hotspots of forest fragmentation, livestock and human density. We also identify areas susceptible to new hotspot emergence in response to moderate expansion of urbanization, livestock production, or forest disturbance, thereby highlighting regions vulnerable to SARS-CoV spillover under future land-use change. In China population growth and increasing meat consumption associated with urbanization and economic development have expanded the footprint of agriculture, leading to human encroachment in wildlife habitat and increased livestock density in areas adjacent to fragmented forests. The reduced distance between horseshoe-bats and humans elevates the risk for SARS-related coronavirus transmission to humans.

**Sentence summarizing manuscript:** Wildlife reservoirs for SARS-coronavirus-2 live in global hotspots of forest fragmentation, livestock, and human density in China

## Introduction

The ongoing severe acute respiratory syndrome coronavirus-2 (SARS-CoV-2) pandemic is claiming human lives and disrupting the functioning of human societies in unprecedented ways. Where did this virus come from and how did it transmit to humans? What facilitated such a host shift and how can we prevent this from happening again? These are important questions, in addition to the most outstanding ones about whether, when, and how this pandemic will end.

Recent years have seen a rise in the recorded number of epidemics from emerging diseases. Such epidemics constitute a major public health threat because of our limited knowledge of prevention and treatment therapies. Most emerging infectious diseases originate from pathogen spillovers from wildlife to humans (1). Why are such outbreaks increasing? Does environmental change play a role?

Crucial to understanding the emergence of infectious diseases is the analysis of the factors facilitating the spillover from wildlife prior to further spread within human populations (2,3). In the case of SARS-CoV-2 genomic sequencing has shown that the virus is most closely related (∼96%) to a strain present in horseshoe bats (4). It is still unclear whether the spillover of SARS-CoV-2 occurred directly from bats to humans or through an intermediate species. For instance, a strain of coronavirus very similar to SARS-CoV-2 was detected in Malayan Pangolin (*Manis javanica*) (5), a wild mammal that is frequently illegally smuggled from Southeast Asia into China and sold in markets (5). Regardless of the specific pathway, the pathogen flow of emerging zoonotic diseases to humans is the result of human interactions with wildlife. We argue that the increasing incidence of emerging disease outbreaks is the result of a similar set of drivers able to change the distance and contact rates between wildlife and humans (as well as human-human interaction), including population growth, urbanization, increasing affluence in mid-income countries, and the associated dietary shifts (6). As countries become more affluent demand for animal products increases, leading to an expansion of agriculture and animal husbandry, often at the expense of natural ecosystems (7). Human penetration in wildlife habitat favors the interaction between humans and wildlife species, either directly through activities like hunting or through other species, particularly livestock that are in closer contact with humans (8-11). The establishment of pastures, plantations or concentrated livestock farms close to forest margins may increase pathogen flows from wildlife to humans (2,9,11-14). Indeed, deforestation and forest fragmentation themselves reshape the dynamics of wildlife communities, possibly leading to the extinction of habitat specialist species, while allowing generalists to thrive (15).

While several reports in the media have conjectured on a possible link between land use change and the emergence of the COVID19 pandemic, such a hypothesis still has to be supported by a comprehensive high-resolution analysis of land use patterns that combines forest fragmentation to livestock and human encroachment in wildlife habitat (2). Here we analyze environmental changes to explain why China remains at risk for other SARS-related coronavirus outbreaks (1,2). We analyze a set of factors that could make China a suitable location for the spillover to humans to occur. To that end, while we do not specifically link environmental change or bats as the immediate hosts of the SARS-CoV-2 ancestor (4,5), we use horseshoe bats in the genus *Rhinolophus* (family Rhinolophidae) as a model system to understand the risk of future coronavirus outbreaks because China is reported to be a region with both highly diverse horseshoe bats and bat SARS-like CoV (16-18).

## Results

Among the four CoV genera, two (alpha- and betacoronaviruses) are found in bats. Among the betacoronaviruses, all four subgenera have been discovered in bats, including the SARS-related CoVs (SARSr-CoV, subgenera *Sarbecovirus*) (16-18). Both SARS-CoV-1 and Swine acute diarrhoea syndrome coronavirus (SADS-CoV) emerged in southeast China and were later detected in horseshoe bats, mainly *Rhinolophus sinicus* and *Rhinolophus affinis* (4,16-18). Most SARSr-CoVs are detected in horseshoe bats, although some strains have been detected in other genera. SARSr-CoVs in China are most similar to the highly pathogenic human SARS-CoVs (4,16-18). Therefore, we performed our local analyses of disturbance at bat locations to horseshoe bats in China (Fig 1a; Table S1) and our analyses of disturbance within horseshoe bat distributions in both the larger South, East, and South East Asia region and then China.

**Figure 1.**
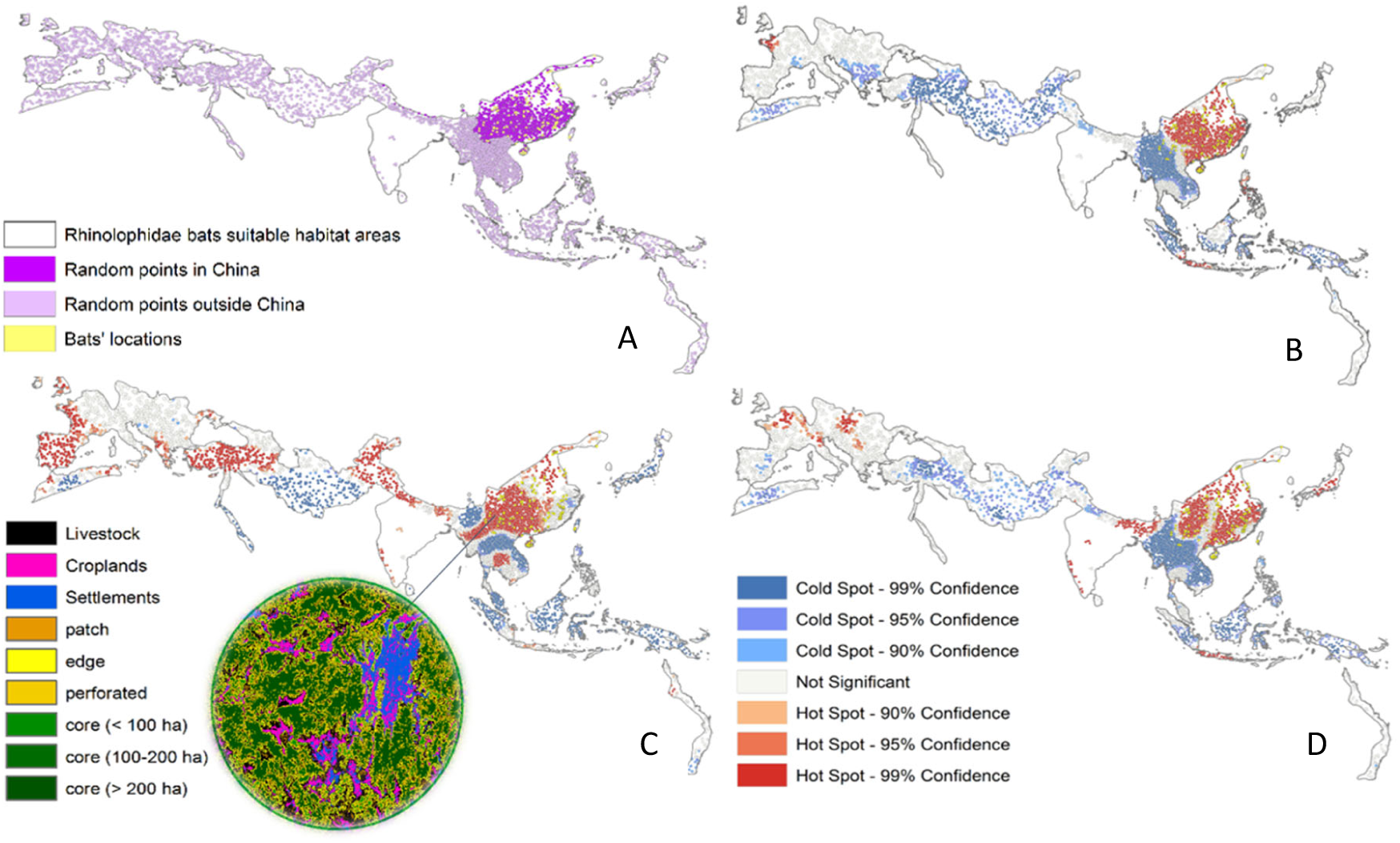
Univariate spatial analysis of coronavirus outbreak drivers (A) Sampling points randomly generated within China (dark purple) and outside China (light purple) and bat location points (yellow), weighted by the horseshoe bat species distributions present in East, South & South East Asia; (B) hotspots (red) and coldspots (blue) of livestock density; (C) hotspots of forest fragmentation; (D) hotspots of human settlement.

Within these distributions we generated 10000 random sampling points (Fig 1a, see SI). Within 30km from every random sampling point we calculate livestock density (n/km^2^), forest cover and fragmentation, cropland cover, population density, and the fractional cover of human settlements (see Supplementary Materials) (14). Hotspots were calculated (Fig 1a) using the Getis-Ord algorithm to show the areas with high or low values of land use attributes cluster (Fig 2).

**Figure 2.**
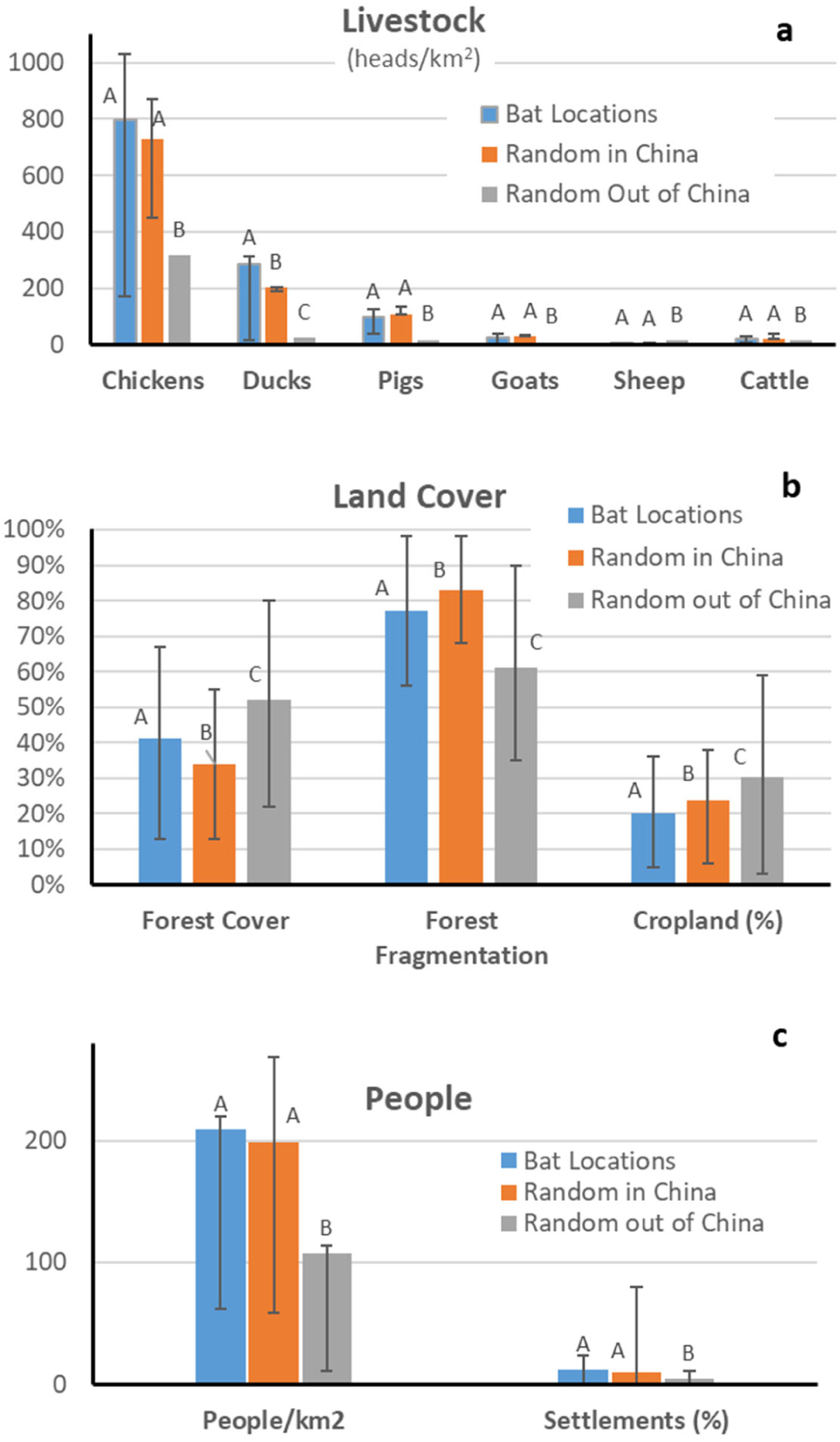
Distribution comparison for coronavirus outbreak drivers. Average distributions of livestock (A), land cover and use (B), and human population (C) in areas likely suitable for horseshoe bat occurrence in China and in the rest of their distribution. Error bars correspond to the 20% and 80% percentiles. Different capital letters indicate statistically different means (α=0.05).

China exhibits a relatively high concentration of livestock production in horseshoe bat distributions (Fig 1a, Table S1). Indeed, China is a hotspot of livestock density within this region (Fig 1b), with statistically significant higher concentrations of chickens, ducks, pigs, goats and cattle (Fig 2a). Within a 30km radius from observed bat locations the density of chicken, ducks, pigs, goats, and cattle was again significantly greater than randomly selected locations. Conversely the sheep density is lower in China, though sheep density was low overall, as it was for other ruminants. The density of chickens, pigs, goats and cattle surrounding (<30km) the points where these bats were recorded and at the randomly selected locations in China within the suitability region were not significantly different, indicating that these random locations have livestock densities that are representative of the areas in which the actual presence of horseshoe bats has been documented.

Forest cover and fragmentation have been related to virus outbreaks from wildlife (including bats) for other zoonotic diseases such as Ebola virus disease (14). China exhibits on average lower forest cover and cropland density and greater forest fragmentation than the other regions analyzed (Fig 1c). The average forest cover and forest fragmentation in the surroundings (within 30km distance) of random points selected in China and the other regions (Figs 1a, 2b) show that these differences are statistically significant. Likewise, statistically significant differences (i.e. lower average cover and higher average fragmentation) are found between the points of actual observations of horseshoe bats and randomly selected locations in the regions outside China within the distributions of these bats (Fig 2b).

China also exhibits higher levels of human presence in horseshoe bat distributions, as evidenced by population density and the fraction of the landscape covered by villages, towns, and other human settlement (Fig 2c). Indeed, the region of China suitable for horseshoe bats coincides with hotspots of human settlements (Fig 1d). Collectively, these results demonstrate that China exhibits stronger signs of human encroachment, livestock density, and forest disturbance of SARSr-CoV hosting horseshoe bat distributions than other regions. In China, regions close to forest fragments are more densely used for livestock production and human settlements – and consequently exhibit lower forest and cropland cover (Fig 1b) – thereby favoring the contact between wildlife and humans either directly or through intermediate animals such as livestock. The fact that China is a global hotspot in the concurrence of these three factors (fragmentation, livestock density, and human settlement) is highlighted by the multivariate hotspot analysis (Fig. 3). These three attributes account for bat habitat (fragmentation), livestock, and human presence, which are major factors contributing to the spillover of zoonotic infectious diseases. Interestingly, we find that China is the global hotspot of simultaneously high forest fragmentation, livestock density and human settlement. The other major global hotspots outside China are found in Java, Bhutan, east Nepal, northern Bangladesh, the Kerala state of India and North-East India, the latter of which are known for past outbreaks of Nipah virus, another bat-related zoonotic disease (19).

**Figure 3.**
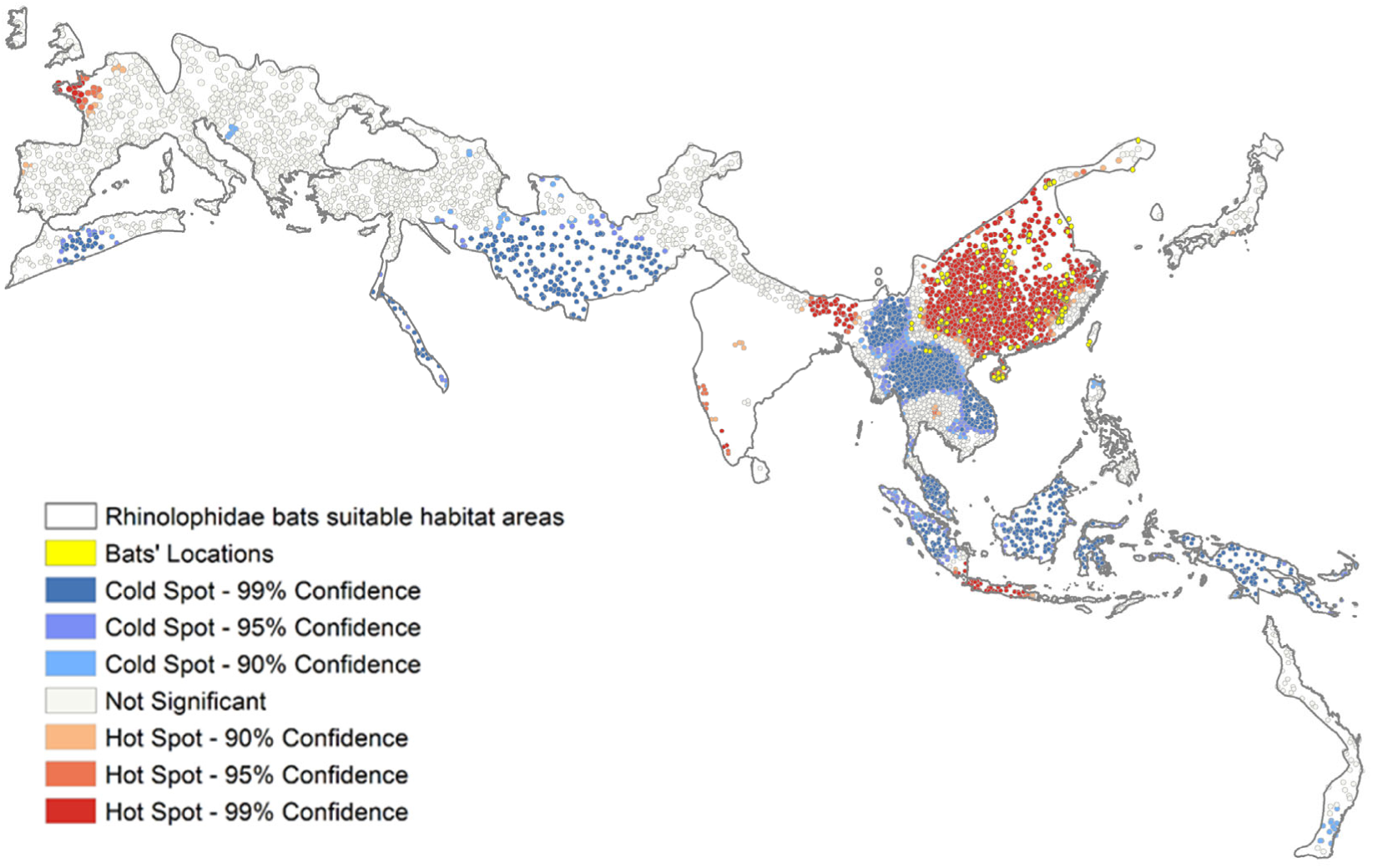
Multivariate spatial analysis of coronavirus outbreak drivers. Hotspot analysis based on the average Gi* Z-Score values for fragmentation, livestock density, and human settlements. Hotspots are classified based on their significance level.

We then use the multivariate hotspot framework to identify regions at high potential risk of SARSr-CoVs spillovers to humans as a result of land use change. To that end the results of the multivariate hotspot analysis were clustered in 30 groups, based on geographic contiguity and similarity in the above three attributes (Fig. 4a; Table S3). We then perturbed one attribute at a time in each group to evaluate that group’s susceptibility to transitioning from non-significant conditions (Fig. 3) to a hotspot state (Fig. 4b). This sensitivity analysis (Fig 4b) shows areas at risk of transitioning to hotspots as a result of a future increase in at least one of the analyzed attributes (i.e. forest fragmentation, livestock density, or human settlement). Interestingly the Chinese region south of Shanghai is at high risk of potentially turning into a hotspot as a result of fragmentation increase. Other regions susceptible to hotspot transition as a result of forest fragmentation include Japan and north Philippines. Likewise, the transition region between China’s hotspot and Indochina’s coldspot and the region surrounding the hotspot of Thailand could turn into hotspots for SARSr-CoV spillover as a result of increasing presence of livestock or humans, respectively (Fig. 4b). These results point both to regions of the world currently suitable for SARSr-CoV spillover from wildlife to humans as well as those at risk of becoming prone to spillover as a result of trajectories of land use change and human penetration (Fig. 4c; Fig.S8 SI)

**Figure 4.**
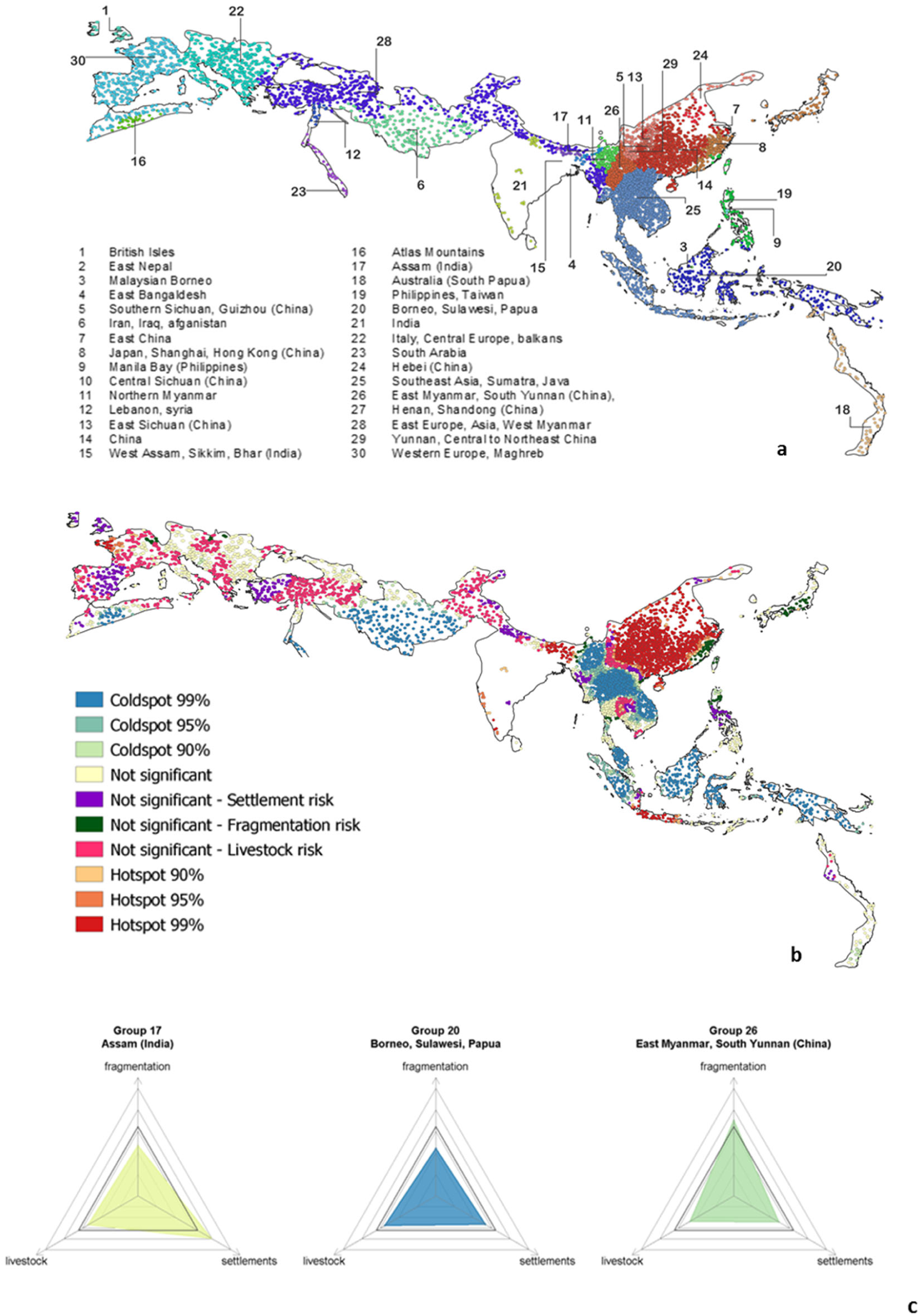
Multivariate grouping analysis of coronavirus spillover drivers. (A) Multivariate groups analysis based on fragmentation, livestock and settlements attributes; (B) Areas at risk of becoming hotspots as a result of changes in forest fragmentation (green), increase in livestock density (fuchsia) and human settlement (purple); (C) Possible trajectories of hotspot transition. These 3 groups represent areas not yet classified as hotspots (*not significant* by Gettis Ord analysis), but which may change trajectory. The solid triangle represents the safe space of variation for the 3 indicators (using multivariate Getis-Ord analysis).

## Discussion

Our approach uses the horseshoe bats as a model family because of their key role as hosts of *Sarbecovirus* coronaviruses, which have caused SARS, COVID-19, and SADS (4,13-18). Other strains of related viruses have been found in other bat genera, but these relationships are less clear (16-18). The widespread sampling of other bats may find species-specific relationships, though horseshoe bats appear to be the reservoirs where most SARSr-CoVs have their evolutionary ancestors so we assume they are the most appropriate model. The risk to humans from other coronaviruses, therefore, will be different, because their host distributions are different, and two CoV genera (gamma- and delta-coronaviruses) are mostly bird viruses.

The bat location data and species distribution data also suffer from different, but related issues. The bat location data are presence only data. True absence data are difficult to obtain, therefore we randomly sampled within different locations to generate pseudo-absence data. Choosing where to sample from also present difficulties, so we chose horseshoe bat distribution data for species that existed within China, East Asia, South, and South East Asia. This presents further issues as the distribution of one species, the greater horseshoe bat (*Rhinolophus ferrumequinum*), extends from Western Europe, Northern Africa, Central Asia and Eastern Asia. We therefore weighted our sampling based on the number of overlapping species distributions to account for this. However, these species distributions are large polygons and the realized niches used within them by the species likely differ, so better niche models using presence and, ideally, presence/absence data are required to develop better species presence predictions (20). However, our results for random locations in China and outside China and reported bat observations were comparable.

More generally, though using the relatively specific bat and virus relationships, we took a high-level approach to understand the more distal or ultimate (rather than proximal) causes of infectious disease emergence in China, linking environmental change and human drivers like agricultural intensification. Different infectious diseases have different transmission mechanisms and life cycles, and not all will respond to such changes in the same way. For example, directly transmitted, acute infections with short incubation and infectious periods, like SARSr-CoVs, will likely be dependent on hosts having greater densities, like in China, for them to emerge. The epidemic potential is then also increased through local and global movement and trade, either of people, wildlife, or livestock (8,21-23). Along with the biological properties of the virus and hosts, the true risk of both the initial cross-species transmission and epidemic potential is either increased or limited by more proximal mechanisms, such as biosecurity, health and safety measures (e.g. personal protective equipment, meat hygiene) that can reduce risk, even if the ultimate factors are present and increasing through the processes of habitat fragmentation and human encroachment (8,14).

Spillover of infectious disease such as SARS, COVID-19 and SADS from wildlife to humans likely requires the coexistence of horseshoe bats and humans in the same environment and is favored by the presence of intermediate animal species, particularly livestock because it is in closer contact with humans. The fragmentation and disturbance of forest ecosystems likely favors habitat generalist bat species. In particular, chickens, ducks, and pigs have been associated with the spread of several zoonotic viral infections, such as influenza viruses. This study demonstrates that in China these important factors responsible for reducing the distance between wildlife and humans co-occur both in horseshoe bat distributions and in the surroundings of actual documented bat occurrence. These results are consistent with the notion that population growth and increasing meat consumption associated with urbanization and economic growth have expanded the footprint of agriculture, leading to human encroachment in wildlife habitat and increased livestock density in areas adjacent to fragmented forest patches. China has dramatically increased animal consumption (24), likely as the result of increasing affluence. In China, meat supply is largely reliant on domestic production using imported feed (e.g. soy from the Americas) (24), which explains the high livestock density in many rural areas, including those at the forest margins. Likewise, economic growth and the shift to diets richer in animal products explains the increasing demand for wild animal meat delicacies, increasing human-wildlife interactions through multiple pathways and the disturbance of forest habitat in more remote locations – frequently abroad – through trade-related teleconnections (25).

The multivariate hotspot analysis highlights how China is the largest hotspot for the concurrence of high forest fragmentation, livestock density, and human presence in our analysis (Fig. 3). The sensitivity analyses identifying the possible transition to new hotspots in response to an increase in one of these attributes (Fig. 4b) highlights areas that could become suitable for spillover and the type of land use change that could induce hotspot activation. Therefore, this analysis highlights region-specific targeted interventions that are urgently needed to increase resilience to SARSr-CoV spillovers. For instance, the green dots in Fig. 4 could be turned into hotspots as a result of forest fragmentation. In these regions resilience can be built through forest conservation or restoration efforts. Indeed, land use change evaluations should consider the risk of activating new hotspots suitable for wildlife-to-human spillover of pathogens such as SARSr-CoV, an aspect that has seldom been included in the impact analysis of land use change. Likewise, other regions such as the China-Indochina transition zone or central Thailand are prone to hotspot transitioning as a result of increased livestock density of urbanization, respectively. In these cases, mitigation of SARSr-CoV emergence can been enhanced by reducing livestock or human density, respectively, thereby inverting ongoing dietary and urbanization trends. Thus, environmental health, is tightly connected to both animal and human health, as recently stressed by planetary and ‘one health’ discourses, which advocate for more holistic views of global health, encompassing environment, animals, and people as well as their interactions (26).

## Materials and Methods

### Bat location data

Most SARS-related CoVs are detected in horseshoe bats, although some strains have also been detected in other genera (22,27-33). SARSr-CoVs in China are most similar to the highly pathogenic human SARS-CoVs (34-36).

We restricted our local analyses of disturbance at bat locations to Rhinolophid bats in China. We performed a Web of Science® search on 10/04/2020 using the following Boolean Operators: Rhinoloph* AND China AND Monitor* OR Survey OR Niche OR Distribution. We found 129 unique references. We removed all those published before 2000, reporting data outside China, review articles, and non-English (specifically 23 Chinese language publication), those with no Rhinolophid data, and those reporting only fossil records. We kept infection studies. This left 48 publications. We then further manually reviewed the publications for those reporting location data (22) but more specifically those with latitude and longitude, leaving 17 publications and 264 observations (see Fig. 1a; Table S1).

### Bat distribution data

We restricted our analyses of disturbance in bat distributions to Rhinolophid bats in both the larger South, East and South East Asia region (but see main manuscript) and then China. We searched the IUCN Red List database (https://www.iucnredlist.org/search) using Taxonomy: Rhinolophidae and Region: East Asia and South & South East Asia (herein ‘regional’) followed by Taxonomy: Rhinolophidae and Region: East Asia: China, Hong Kong & Taiwan (herein ‘Chinese’) classifications and downloaded the shapefiles for the 55 regional and 22 Chinese Rhinolophus species present in the region. We consider these areas as regions of suitable habitat for Rhinolophidae. The extent of this study area exceeds 28.5 million km^2^.

Within these putative species distributions, we generated 10000 random sampling points with a local sampling density that is proportional to the number of species whose distributions were reported at the point. For every random sampling point we consider a circular area of 30km radius within which we calculate livestock density, forest cover and fragmentation, cropland cover, population density and the fractional cover of human settlements as explained below. The average values of these statistics are then calculated for China and the other regions of the world with habitat suitable for Rhinolophidae and compared and the difference is tested for significance using the Mann-Whitney non-parametric test in Mathematica©.

### Livestock, forest cover, and population data

We took livestock data from the GeoWiki database that provides georeferenced livestock counts (in heads/km^2^) at 1 km resolution for chickens, ducks, pigs, goats, sheep, and cattle (https://livestock.geo-wiki.org/home-2/). We quantified human presence both in terms of population density at 1 km resolution and as fraction of the landscape taken by villages, towns or other settlements from the WorldPop database at 1 km resolution. We used cropland data (at 30×30m^2^ resolution) from (37). Forest cover data are available at 30m resolution annually between 2000 and 2018 (38). Forest cover is associated with the presence of trees taller than 5 m. Forest loss or gain was determined as the difference in forest cover between these two years.

### Forest fragmentation analyses

We performed a forest fragmentation analyses based on Vogt et al. (39) using the 30m forest cover data. This method distinguishes forest cores, from forest margins and patches. Every 30m pixel is classified as wooded or non-wooded, based on whether its tree cover was greater or smaller than 50% in the year 2018. Forest cores are wooded pixels that are not adjacent to non-wooded pixels. Conversely, forest patches are made of wooded pixels that are not adjacent to forest core pixels. Wooded pixels that are neither core nor patch pixels occur at the margins of forest cores. Forest fragmentation was then quantified in terms of a composite fragmentation index (CFI) (13), defined as the ratio between the sum of number of pixels classified as “margins”, “patches”, or smaller core areas (i.e.,<200 ha), and the total number of pixels (wooded + non-wooded) in the 30km circles used to characterize land cover and land use in the surroundings of the points of actual bat observations or the randomly generated points. This index ranges between 0 and 1.

### Hotspot Analyses and multivariate clustering

We mapped statistically significant hotspots of livestock density, forest fragmentation, and human settlements. To be a hotspot, an area needs to have a high (or low, in the case of cold-spots) value of its attribute and be also surrounded by other areas with high (or low) values of the same attribute. To that end, the 30km circles centered on the 10000 random points (Fig. 1a) considered in this study were used to carry out a hotspot analysis applying the Getis-Ord algorithm (Gi* statistics). The Gi* statistic shows to what extent areas with high or low values tend to cluster thereby forming a hotspot. The result of the Gi* analysis is reported in terms of statistically significance with 90%, 95%, and 99% confidence (Fig. 1).

We then used two different methods to generate a multivariate distribution for the three indicators (livestock density, forest fragmentation and human settlements). First, we averaged their Gi*. Since the Gi* is a z-score, i.e. it has a standard normal distribution, a linear combination of the three Gi* indicators, such as their average, is a standard normal distribution and can still be represented with the same significance levels (Fig. 2). Second, we performed a spatially constrained multivariate clustering analysis. A Minimum Spanning Tree (MST) from the connectivity graph of the features was built, and then the SKATER (Spatial “K”luster Analysis by Tree Edge Removal) clustering method was used (40). The SKATER iteratively cuts branches in the MST, based on data variability among and within groups and on a spatial constraint, until it reaches the user-defined number of groups. The spatial constraint defined here is a k nearest neighbors type with 8 neighbors, meaning each feature in a group must have at least one of its 8 nearest neighbors in the same group. We chose 30 as the number of groups, calculated a set of summary statistics and boxplots for the groups and compared them to their global values (Table S2). For each indicator, we calculated the R^2^ value as the reduction in variance of the indicator obtained by grouping, divided by the original variance of the indicator (Table S2). While the modularity analysis based on pseudo F-statistics shows that the optimal number of groups (the maximum differences between groups while maximizing within group similarity) is 12, here we studied 30 groups to analyse distinct regional patterns. Having a greater number of groups allows us to identify groups that are susceptible to transitioning to a hotspot because they are not “too different” from hotspots.

## Data Availability

All data are available

## Acknowledgments

M.C.R and N.G. are supported by Cariplo Foundation (SusFeed project 0737 CUP D49H170000300007). D.T.H. is supported by USDA Hatch Multistate project no. W4190 capacity fund; Rutherford Discovery Fellowship RDF-MAU1701.

## Competing interest statement

The authors declare no competing interests.

## Author contributions

M.C.R. designed research; M.C.R. and D.T.S.H performed research; M.C.R., P.D. and N.G. analyzed data; P.D., M.C.R. and D.T.S.H wrote the paper.

## Supplementary Materials

### Environmental Change and Coronavirus Emergence Risk

### Figures and Tables Supplementary Materials (SI)

**Table S1.** Horseshoe bat locations based on literature survey of studies reporting occurrences and related coordinates (coordinates are here listed as reported in the original articles and therefore they are not in a homogeneous format).

| Species | Site | Location | Region | Latitude | Longitude | Ref. |
| --- | --- | --- | --- | --- | --- | --- |
| *R. macrotis* | Shiyan Cave | Jing gang shan Nature Reserve | Jiangxi Province | 26°36'N | 114°12'E | <sup>1</sup> |
| *R. macrotis* | Xianren Cave | Jinning County | Yunnan Province | 24°30'N | 102°20'E |  |
| *R. macrotis* | Tiantang Cave | Changtang County, Nanning city | Guangxi Province | 22°49'N | 108°42'E |  |
| *R. luctus* | Emei shan | Emei shan | Sichuan province | 29°31'11"N | 103°19'57"E | <sup>2</sup> |
| *R. formosae* | Pingtung | Pingtung | Taiwan | 22°40'16"N | 120°29'17"E |  |
| *R. formosae* | Kaohsiung | Kaohsiung | Taiwan | 23°01'18"N | 120°39'25"E |  |
| *R. affinis* | Shiyan cave | Liping village, Jinggangshan Natural Reserve | Jiangxi Province | 26°36'N | 114°12'E | <sup>3</sup> |
| *R. pearsoni* | Shiyan cave | Liping village, Jinggangshan Natural Reserve | Jiangxi Province | 26°36'N | 114°12'E |  |
| *R. macrotis* | Longxu cave | Shuanghe township | Yunnan Province | 24°30'N | 102°20'E | <sup>4</sup> |
| *R. lepidus* | Longxu cave | Shuanghe township | Yunnan Province | 24°30'N | 102°20'E |  |
| *R. sinisus* | Longxu cave | Shuanghe township | Yunnan Province | 24°30'N | 102°20'E |  |
| *R. pusillus* | Longxu cave | Shuanghe township | Yunnan Province | 24°30'N | 102°20'E |  |
| *R. ferrumequinum* | Longxu cave | Shuanghe township | Yunnan Province | 24°30'N | 102°20'E |  |
| *R. rex* | Longxu cave | Shuanghe township | Yunnan Province | 24°30'N | 102°20'E |  |
| *R. rex* | Qianling Park | Guiyang | Guizhou province | 26°36'58.0"N | 106°41'26.6"E | <sup>5</sup> |
| *R. ferrumequinum* | Dalazi Cave | Zhi'an village, Ji'an city | Jilin Province | 41°3'55.8"N | 125°50'9.8"E | <sup>6</sup> |
| *R. affinis* |  | Jing Xi County Nature Preserve | Guangxi Autonomous Region | 23°07'12" N | 105°57'36" E | <sup>7</sup> |
| *R. affinis* |  | Shiwandashan National Nature Preserve | Guangxi Autonomous Region | 21°13'48" N | 107°52'48" E |  |
| *R. pearsonii* |  | Dashahe Nature Preserve | Guizhou Province | 29°10'12" N | 107°34'12" E |  |
| *R. rouxii* |  | Shiwandashan National Nature Preserve | Guangxi Autonomous Region | 21°13'48" N | 107°52'48" E |  |
| *R. rouxii* | Shuipu | Maolan National Nature Preserve | Guizhou Province | 25°29'05" N | 107°52'54" E |  |
| *R. ferrumequinum* |  | Huizhe | Yunnan | 26:28:53 | 103:27:06 | <sup>8</sup> |
| *R. ferrumequinum* |  | Baoxing | Sichuan | 30:32:57 | 102:42:38 |  |
| *R. ferrumequinum* |  | Emei Shan | Sichuan | 29:34:74 | 103:16:92 |  |
| *R. ferrumequinum* |  | Nanton | Sichuan | 29:48:33 | 102:13:52 |  |
| *R. ferrumequinum* |  | Baiya | Shaanxi | 33:21:32 | 108:03:36 |  |
| *R. ferrumequinum* |  | Foping | Shaanxi | 33:34:00 | 107:59:00 |  |
| *R. ferrumequinum* |  | Shuangtai | Hubei | 32:28:20 | 110:13:32 |  |
| *R. ferrumequinum* |  | Shuidong | Hubei | 32:57:56 | 110:25:06 |  |
| *R. ferrumequinum* |  | Tiangou | Hubei | 32:25:24 | 110:08:50 |  |
| *R. ferrumequinum* |  | Shenxian | Henan | 34:41:49 | 113:22:05 |  |
| *R. ferrumequinum* |  | Shuanglongwan | Henan | 33:58:28 | 110:56:57 |  |
| *R. ferrumequinum* |  | Yingyang | Henan | 34:01:19 | 113:14:40 |  |
| *R. ferrumequinum* |  | Zhangjiashan | Henan | 34:28:29 | 110:48:19 |  |
| *R. ferrumequinum* |  | Jindezhen | Jiangxi | 29:19:44 | 117:11:48 |  |
| *R. ferrumequinum* |  | Wanfu | Jiangxi | 28:05:47 | 117:01:46 |  |
| *R. ferrumequinum* |  | Jinzhai | Anhui | 31:25:05 | 115:44:44 |  |
| *R. ferrumequinum* |  | Youkuang | Anhui | 31:10:01 | 115:19:59 |  |
| *R. ferrumequinum* |  | Changshan | Shandong | 34:59:37 | 117:58:44 |  |
| *R. ferrumequinum* |  | Nanzhi | Shandong | 36:00:08 | 117:00:04 |  |
| *R. ferrumequinum* |  | Xiaya | Shandong | 36:15:00 | 118:07:01 |  |
| *R. ferrumequinum* |  | Yinan | Shandong | 35:31:06 | 118:24:40 |  |
| *R. ferrumequinum* |  | Beijing | Beijing | 39:54:25 | 116:19:55 |  |
| *R. ferrumequinum* |  | Fangshan | Beijing | 39:45:54 | 115:58:56 |  |
| *R. ferrumequinum* |  | Jietai | Beijing | 39:54:35 | 116:14:55 |  |
| *R. ferrumequinum* |  | Jilin Shi | Jilin | 43:46:49 | 126:29:56 |  |
| *R. marshalli* | Tiantang Cave | Tiantang village, Nanning city | Guangxi Province | 4183955.80 N | 12585099.80 E | <sup>9</sup> |
| *R. ferrumequinum* |  | Emei Shan | Sichuan | 29°34'74"N | 103°16'92"E | <sup>10</sup> |
| *R. ferrumequinum* |  | Foping | Shaanxi | 33°34'00"N | 107°59'00"E |  |
| *R. ferrumequinum* |  | Daguping | Shaanxi | 33°34'00"N | 107°46'00"E |  |
| *R. ferrumequinum* |  | Beijing | Hebei | 39°54'25"N | 116°19'55"E |  |
| *R. ferrumequinum* | Fish cave |  | Anhui Province | 30u209N | 117u509E | <sup>11</sup> |
| *R. sinicus* |  | Qingyang, Anhui | Qingyang, Anhui | N30:20:511 | E117:50:128 | <sup>12</sup> |
| *R. sinicus* |  | Jingxian, Anhui | Jingxian, Anhui | N30:26:785 | E118:24:783 |  |
| *R. sinicus* |  | Huashanmiku, Anhui | Huashanmiku, Anhui | N29:45:199 | E118:23:331 |  |
| *R. sinicus* |  | Sanling mountain, Jiangxi | Sanling mountain, Jiangxi | N29:22:112 | E117:34:324 |  |
| *R. sinicus* |  | Qingfeng cave, Jiangxi | Qingfeng cave, Jiangxi | N29:22:262 | E117:39:357 |  |
| *R. sinicus* |  | Qinhui cave, Jiangxi | Qinhui cave, Jiangxi | N29:22:662 | E117:32:335 |  |
| *R. sinicus* |  | Longhu mountain, Jiangxi | Longhu mountain, Jiangxi | N28:04:107 | E116:58:227 |  |
| *R. sinicus* |  | Lijia country, Jiangxi | Lijia country, Jiangxi | N28:06:726 | E116:59:282 |  |
| *R. sinicus* |  | Wuyishan baohuqu, Fujian | Wuyishan baohuqu, Fujian | N27:44:000 | E117:40:000 |  |
| *R. sinicus* |  | Wuyishan tiliqiao, Fujian | Wuyishan tiliqiao, Fujian | N27:44:543 | E117:29:953 |  |
| *R. sinicus* |  | Wuyishan Yanzijiao, Fujian | Wuyishan Yanzijiao, Fujian | N27:48:511 | E117:42:505 |  |
| *R. sinicus* |  | Taihe, Jiangxi province | Taihe, Jiangxi province | N26:36:151 | E114:12:734 |  |
| *R. sinicus* |  | Jinggang mountain, Jiangxi | Jinggang mountain, Jiangxi | N26:31:215 | E115:06:610 |  |
| *R. sinicus* |  | Xingguo, Jiangxi | Xingguo, Jiangxi | N26:19:314 | E115:35:229 |  |
| *R. sinicus* |  | Taining, Fujian | Taining, Fujian | N26:42:236 | E117:29:867 |  |
| *R. sinicus* |  | Jiangle, Fujian | Jiangle, Fujian | N26:39:537 | E117:34:387 |  |
| *R. sinicus* |  | Mingxi, Fujian | Mingxi, Fujian | N26:21:440 | E117:11:398 |  |
| *R. sinicus* |  | Yongan, Fujian | Yongan, Fujian | N25:51:500 | E117:17:000 |  |
| *R. sinicus* |  | Liancheng, Fujian | Liancheng, Fujian | N25:12:404 | E117:15:066 |  |
| *R. sinicus* |  | Shanghang, Fujian | Shanghang, Fujian | N25:15:020 | E116:49:009 |  |
| *R. sinicus* |  | Guilin, Guangxi | Guilin, Guangxi | N25:16:278 | E110:17:009 |  |
| *R. sinicus* |  | Ruyuan, Guangdong | Ruyuan, Guangdong | N24:59:086 | E113:08:523 |  |
| *R. sinicus* |  | Luofushan, Guangdong | Luofushan, Guangdong | N23:15:589 | E114:03:656 |  |
| *R. sinicus* |  | Zhangjiajie, Hunan | Zhangjiajie, Hunan | N29:21:410 | E110:34:783 |  |
| *R. sinicus* |  | Yongshun, Hunan | Yongshun, Hunan | N29:03:720 | E109:38:358 |  |
| *R. sinicus* |  | Jishou, Hunan | Jishou, Hunan | N28:18:208 | E109:39:175 |  |
| *R. sinicus* |  | Fenghuang, Hunan | Fenghuang, Hunan | N27:59:580 | E109:33:786 |  |
| *R. sinicus* |  | Wuchuan, Guizhou | Wuchuan, Guizhou | N28:34:237 | E107:54:058 |  |
| *R. sinicus* |  | Anlong, Guizhou | Anlong, Guizhou | N25:16:577 | E105:31:931 |  |
| *R. sinicus* |  | Emeishan, Sichuan | Emeishan, Sichuan | N29:34:803 | E103:24:708 |  |
| *R. sinicus* |  | Huize, Yunnan | Huize, Yunnan | N26:42:000 | E103:30:000 |  |
| *R. sinicus* |  | Jiuxiang, Yunnan | Jiuxiang, Yunnan | N25:07:000 | E103:22:000 |  |
| *R. sinicus* |  | Fumin, Yunnan | Fumin, Yunnan | N25:11:796 | E102:27:863 |  |
| *R. sinicus* |  | Yongde, Yunnan | Yongde, Yunnan | N24:21:427 | E099:02:161 |  |
| *R. sinicus* |  | Yinggeling, Hainan | Yinggeling, Hainan | N19:04:982 | E109:33:107 |  |
| *R. sinicus* |  | Wuzhishan, Hainan | Wuzhishan, Hainan | N18:46:309 | E109:31:012 |  |
| *R. sinicus* |  | Qiongzong, Hainan | Qiongzong, Hainan | N18:49:589 | E110:00:435 |  |
| *R. sinicus* |  | Baoqing, Hainan | Baoqing, Hainan | N18:42:237 | E109:41:528 |  |
| *R. sinicus* |  | Lingshui, Hainan | Lingshui, Hainan | N18:38:586 | E109:57:636 |  |
| *R. sinicus* |  | Jianfengling, Hainan | Jianfengling, Hainan | N18:47:275 | E108:57:364 |  |
| *R. sinicus* |  | Maogan, Hainan | Maogan, Hainan | N18:36:306 | E109:26:776 |  |
| *R. affinis* | Shiyan cave | Liping village, Jinggangshan Natural Reserve | Jiangxi Province | 26°36'N | 114°12'E | 13 |
| *R. pearsoni* | Shiyan cave | Liping village, Jinggangshan Natural Reserve | Jiangxi Province | 26°36'N | 114°12'E |  |
| *R. macrotis* |  | Yangchun | Guangdong Province | 111.94 | 22.44 | 14 |
| *R. macrotis* |  | Shaoguan | Guangdong Province | 113.56 | 24.77 |  |
| *R. macrotis* |  | Nanning | Guangxi Province | 108.70 | 22.82 |  |
| *R. macrotis* |  | Lengshuijiang | Hunan Province | 111.57 | 27.75 |  |
| *R. macrotis* |  | Jianggangshan | Jiangxi Province | 114.20 | 26.60 |  |
| *R. macrotis* |  | Jinning | Jinning | 102.33 | 24.50 |  |
| *R. macrotis* |  | Yuanjiang | Yunnan Province | 102.00 | 23.59 |  |
| *R. macrotis* |  | Baoshan | Yunnan Province | 99.10 | 25.08 |  |
| *R. macrotis* |  | Wulong | Chongqing City | 107.95 | 29.27 |  |
| *R. macrotis* |  | Yangchun | Guangdong Province | 111.94 | 22.44 |  |
| *R. macrotis* |  | Xishui | Guizhou Province | 106.21 | 28.45 |  |
| *R. macrotis* |  | Dazhou | Sichuan Province | 107.74 | 31.20 |  |
| *R. macrotis* |  | Hanzhong | Shannxi Province | 107.03 | 32.84 |  |
| *R. macrotis* |  | Xichuan | Henan Province | 111.55 | 32.87 |  |
| *R. macrotis* |  | Wulong | Chongqing City | 107.95 | 29.27 |  |
| *R. macrotis* |  | Lengshuijiang | Hunan Province | 111.57 | 27.75 |  |
| *R. macrotis* |  | Jianggangshan | Jiangxi Province | 114.20 | 26.60 |  |
| *R. macrotis* |  | Ganzhou | Jiangxi Province | 114.09 | 25.46 |  |
| *R. macrotis* |  | Hanzhong | Shannxi Province | 107.03 | 32.84 |  |
| *R. sinicus* |  | 10km Mojiang, Zhenyuan, and Ning'er towns, Mojiang | Yunnan Province |  |  | 15 |
| *R. affinis* |  | 10km Mojiang, Zhenyuan, and Ning'er towns, Mojiang | Yunnan Province |  |  |  |
| *R. schnitzleri* | Xiao-dong Cave | Gengjiaying Commune, Yiliang County, Kunming City | Yunnan province | 25°02'N | 103°14'E | 16 |
| *R. affinis* | Longkong Cave | Yanshi County | Fujian | 25°12'N | 117°15'E | 17 |
| *R. affinis* | Zhuxi Cave | Shanghang County | Fujian | 25°10'N | 116°45'E |  |
| *R. affinis* | Jinkuang Cave | Liancheng County | Fujian | 25°39'N | 116°53'E |  |
| *R. affinis* | Qixian Cave | Sha County | Fujian | 26°25'N | 117°39'E |  |
| *R. affinis* | Guixian Cave | Mingxi County | Fujian | 26°24'N | 117°11'E |  |
| *R. affinis* | Huanghuacong Cave | Yanzijiao Town, Wuyi Mountain | Fujian | 27°49'N | 117°43'E |  |
| *R. affinis* | Shui Cave | Longmen County | Guangdong |  |  |  |
| *R. affinis* | Dongbian Cave | Longmen County | Guangdong |  |  |  |
| *R. affinis* | Penglai Cave | Yunfu Mount, Bolo County | Guangdong |  |  |  |
| *R. affinis* | Fangkong | Luofu Mountain, Huizhou City | Guangdong | 23°16'N | 114°04'E |  |
| *R. affinis* | Seven Star Cave | Guilin City | Guangxi | 25°17'N | 110°18'E |  |
| *R. affinis* | Fenkeng Cave | Guilin City | Guangxi | 25°17'N | 110°21'E |  |
| *R. affinis* | Seven Star Cave | Guilin City | Guangxi | 25°17'N | 110°18'E |  |
| *R. affinis* | Bianfu Cave | Lingshui County | Hainan | 18°42'N | 109°53'E |  |
| *R. affinis* | Xiashui Cave | Qiongzhong County | Hainan | 19°04'N |  |  |
| *R. affinis* | Qianlong Cave | Baoting County | Hainan | 18°34'N | 109°26'E |  |
| *R. affinis* | Fangkong Cave | Yingge Mountain | Hainan | 19°04'N | 109°33'E |  |
| *R. affinis* | Qishier Cave | Huoshan Mountain, Haikou City | Hainan | 19°57'N | 110°12'E |  |
| *R. affinis* | Bianfu Cave | Xixia countryside | Henan |  |  |  |
| *R. affinis* | Longhushan Park | Yingtian City | Jiangxi | 28°04'N | 116°58'E |  |
| *R. affinis* | Jinuo Town | Xishuang banna | Yunnan | 21°58'N | 100°49'E |  |
| *R. affinis* |  | Manfa Village, Xishuang banna | Yunnan |  |  |  |
| *R. affinis* | Dashu Cave | Xishuangbanna | Yunnan |  |  |  |
| *R. affinis* | Jiuxiang Cave | Yiliang County | Yunnan |  |  |  |
| *R. affinis* | Shaft Cave, near Bailong Cave | Mile County | Yunnan | 24°11'N | 103°21'E |  |
| *R. ferrumequinum* | Bianfu Cave | SW of Beijing | Beijing | 39°43'N | 115°43'E |  |
| *R. ferrumequinum* | Shidu Caves | Fangshan District | Beijing | 39°25'N | 115°17'E |  |
| *R. ferrumequinum* | Bianfu Cave | SW of Beijing | Beijing | 39°43'N | 115°43'E |  |
| *R. ferrumequinum* | Bianfu Cave, Beijing | Bianfu Cave, Beijing | Beijing |  |  |  |
| *R. ferrumequinum* | Guanyin Cave, Jietai Temple | Fang shan Town | Beijing | 39°48'N | 115°48'E |  |
| *R. ferrumequinum* | Seven Star Cave | Guilin City | Guangxi | 25°17'N | 110°18'E |  |
| *R. ferrumequinum* | Shenxian Cave | Huancuigu, Rongyang County | Henan | 34°38'N | 113°15'E |  |
| *R. ferrumequinum* |  | Shennongjia | Henan | 31°30'N | 110°23'E |  |
| *R. ferrumequinum* | Sihe Village | Langhe Town, Wudang Mountain | Henan |  |  |  |
| *R. ferrumequinum* | Tong mu Village | Muyu Town, Tanjiawan County | Henan | 31°28'N | 110°25'E |  |
| *R. ferrumequinum* | Wanfu Cave | Jing dezhen County | Jiangxi | 28°37'N | 116°28'E |  |
| *R. ferrumequinum* | Juda Lizi Cave | Zian Village, Jian City | Jilin | 41°04'N | 125°50'E |  |
| *R. ferrumequinum* | Xiaya Cave | Yiyuan County | Shandong |  |  |  |
| *R. ferrumequinum* |  | Nantou Village, Luding County | Sichuan | 29°48'N | 102°14'E |  |
| *R. ferrumequinum* | Dahu Cave | Emeishan | Sichuan | 29°35'N | 103°17'E |  |
| *R. ferrumequinum* | Du Cave | Panshan Mountain, Tianjin City | Tianjin | 25°33'N | 116°60'E |  |
| *R. ferrumequinum* | Heshang Cave | Kunming, Fumin County | Yunnan | 25°12'N | 102°28'E |  |
| *R. ferrumequinum* | Xiaogou Cave | Kunming City | Yunnan | 25°04'N | 103°23'E |  |
| *R. ferrumequinum* | Xiao Cave | Yiliang County | Yunnan |  |  |  |
| *R. lepidus* | Tianlong Cave | Dali County | Yunnan | 25°55'N | 100°05'E |  |
| *R. lepidus* | Xiyu Cave | Fumin County | Yunnan | 25°09'N | 102°39'E |  |
| *R. osgoodi* | Tianlong Cave | Dali County | Yunnan | 25°55'N | 100°05'E |  |
| *R. osgoodi* | Xiyu Cave | Fumin County | Yunnan | 25°09'N | 102°39'E |  |
| *R. luctus* | Jiuba Cave | Jiangle County | Fujian | 26°43'N | 117°29'E |  |
| *R. luctus* | Kangtou Cave |  | Fujian | 26°40'N | 117°36'E |  |
| *R. luctus* | Xianren Cave | Shishan Town, Haikou City | Hainan | 19°44'N | 109°37'E |  |
| *R. luctus* | Tangle Cave | Jishou | Hunan | 28°18'N | 109°39'E |  |
| *R. macrotis* | Shidu Caves | Beijing | Beijing |  |  |  |
| *R. macrotis* | Closed Cave | Ziyuan County | Guangxi |  |  |  |
| *R. macrotis* | Yuanfeng Cave | Qilai County | Sichuan | 30°29'N | 103°24'E |  |
| *R. siamensis* | Yinhua Cave | Jiangle County | Fujian | 26°39'N | 117°34'E |  |
| *R. siamensis* | Zhuxi Cave | Shanghang County | Fujian | 25°10'N | 116°45'E |  |
| *R. siamensis* | Ganru Cave | Liancheng County | Fujian |  |  |  |
| *R. siamensis* | Yinhua Cave | Jiangle County | Fujian | 26°39'N | 117°34'E |  |
| *R. siamensis* | Seven Star Cave | Guilin City | Guangxi | 25°17'N | 110°18'E |  |
| *R. siamensis* | Closed Cave | Ziyuan County | Guangxi |  |  |  |
| *R. siamensis* | Wolong Cave | Xingyi County | Guizhou | 24°9'N | 104°53'E |  |
| *R. siamensis* | Shanjia Cave | Anlong County | Guizhou | 25°19'N | 105°05'E |  |
| *R. siamensis* | Wanfu Cave | Jing dezhen County | Jiangxi | 28°37'N | 116°28'E |  |
| *R. siamensis* |  | Manfa Village, Xishuang banna | Yunnan |  |  |  |
| *R. siamensis* | Heshang Cave | Kunming, Fumin County | Yunnan | 25°12'N | 102°28'E |  |
| *R. marshalli* | Baisha Cave | Guiping County | Guangxi | 23°14'N | 109°54'E |  |
| *R. marshalli* | Shepo Cave | Guiping County | Guangxi | 23°25'N | 110°15'E |  |
| *R. marshalli* | Bailong Cave | Mile County | Yunnan | 24°12'N | 103°14'E |  |
| *R. marshalli* | Gulong Cave | Yuanjiang County | Yunnan | 23°35'N | 101°58'E |  |
| *R. pearsonii* | Yulong Cave | Qingyang County | Anhui | 30°21'N | 117°50'E |  |
| *R. pearsonii* | Longkong Cave | Yanshi County | Fujian | 25°12'N | 117°15'E |  |
| *R. pearsonii* | Yuhua Cave | Jiangle County | Fujian | 26°39'N | 117°35'E |  |
| *R. pearsonii* | Bianfu Cave | Taining City | Fujian | 26°42'N | 117°30'E |  |
| *R. pearsonii* | Ganru Cave | Liancheng County | Fujian |  |  |  |
| *R. pearsonii* | Guwang Cave | Wuyi City | Fujian | 27°42'N | 117°42'E |  |
| *R. pearsonii* | Huanghuacong Cave | Yanzijiao Town, Wuyi Mountain | Fujian | 27°49'N | 117°43'E |  |
| *R. pearsonii* | Shang Cave | Dafu Village, Meihua Town, Lechang County | Guangdong | 25°09'N | 113°04'E |  |
| *R. pearsonii* | Seven Star Cave | Guilin City | Guangxi | 25°17'N | 110°18'E |  |
| *R. pearsonii* | Closed Cave | Ziyuan County | Guangxi |  |  |  |
| *R. pearsonii* |  | Liangjiang Town, Wuming County | Guangxi |  |  |  |
| *R. pearsonii* | Xipo Cave | Wuchuan County | Guizhou |  |  |  |
| *R. pearsonii* | Wanfu Cave | Jing dezhen County | Jiangxi | 28°37'N | 116°28'E |  |
| *R. pearsonii* | Bao Zi Cave | Tian Quan County | Sichuan | 30°10'N | 102°52'E |  |
| *R. pearsonii* | Fufeng Cave | Emeishan | Sichuan | 29°34'N | 103°25'E |  |
| *R. pearsonii* | Jiulao Cave | Emeishan | Sichuan | 29°33'N | 109°47'E |  |
| *R. pearsonii* | Mingjiu Cave | Mongzi County | Yunnan |  |  |  |
| *R. pearsonii* | Pingbian Village | Mongzi County | Yunnan |  |  |  |
| *R. pearsonii* | Wangzhangya Cave | Dawei County | Yunnan |  |  |  |
| *R. pearsonii* | Bianfu Cave | Mengla County | Yunnan | 21°53'N | 101°18'E |  |
| *R. yunnanensis* | Yulong Cave | Qingyang County | Anhui | 30°21'N | 117°50'E |  |
| *R. yunnanensis* | Longkong Cave | Yanshi County | Fujian | 25°12'N | 117°15'E |  |
| *R. yunnanensis* | Yuhua Cave | Jiangle County | Fujian | 26°39'N | 117°35'E |  |
| *R. yunnanensis* | Bianfu Cave | Taining City | Fujian | 26°42'N | 117°30'E |  |
| *R. yunnanensis* | Ganru Cave | Liancheng County | Fujian |  |  |  |
| *R. yunnanensis* | Guwang Cave | Wuyi City | Fujian | 27°42'N | 117°42'E |  |
| *R. yunnanensis* | Huanghuacong Cave | Yanzijiao Town, Wuyi Mountain | Fujian | 27°49'N | 117°43'E |  |
| *R. yunnanensis* | Shang Cave | Dafu Village, Meihua Town, Lechang County | Guangdong | 25°09'N | 113°04'E |  |
| *R. yunnanensis* | Seven Star Cave | Guilin City | Guangxi | 25°17'N | 110°18'E |  |
| *R. yunnanensis* | Closed Cave | Ziyuan County | Guangxi |  |  |  |
| *R. yunnanensis* |  | Liangjiang Town, Wuming County | Guangxi |  |  |  |
| *R. yunnanensis* | Xipo Cave | Wuchuan County | Guizhou |  |  |  |
| *R. yunnanensis* | Wanfu Cave | Jing dezhen County | Jiangxi | 28°37'N | 116°28'E |  |
| *R. yunnanensis* | Bao Zi Cave | Tian Quan County | Sichuan | 30°10'N | 102°52'E |  |
| *R. yunnanensis* | Fufeng Cave | Emeishan | Sichuan | 29°34'N | 103°25'E |  |
| *R. yunnanensis* | Jiulao Cave | Emeishan | Sichuan | 29°33'N | 109°47'E |  |
| *R. yunnanensis* | Mingjiu Cave | Mongzi County | Yunnan |  |  |  |
| *R. yunnanensis* | Pingbian Village | Mongzi County | Yunnan |  |  |  |
| *R. yunnanensis* | Wangzhangya Cave | Dawei County | Yunnan |  |  |  |
| *R. yunnanensis* | Bianfu Cave | Mengla County | Yunnan | 21°53'N | 101°18'E |  |
| *R. rex* | Dongmen Cave | Fushui County | Guangxi |  |  |  |
| *R. rex* | White Dragon Cave | Guilin City | Guangxi |  |  |  |
| *R. rex* | Fenkeng Cave | Guilin City | Guangxi | 25°17'N | 110°21'E |  |
| *R. rex* | Fenkeng Cave | Guilin City | Guangxi | 25°17'N | 110°21'E |  |
| *R. rex* | Xipo Cave | Wuchuan County | Guizhou |  |  |  |
| *R. rex* | Qingxuan Cave |  | Yunnan | 26°40'N | 100°12'E |  |
| *R. rex* | Tianlong Cave | Dali County | Yunnan | 25°55'N | 100°05'E |  |
| *R. rex* | Bianfu Cave | Yiliang County | Yunnan | 25°04'N | 103°23'E |  |
| *R. rex* | Xiyou Cave | Fumin County | Yunnan | 25°09'N | 102°39'E |  |
| *R. rex* | Xiaogou Cave | Yiliang County | Yunnan |  |  |  |
| *R. paradoxolophus* | Dongmen Cave | Fushui County | Guangxi |  |  |  |
| *R. paradoxolophus* | White Dragon Cave | Guilin City | Guangxi |  |  |  |
| *R. paradoxolophus* | Fenkeng Cave | Guilin City | Guangxi | 25°17'N | 110°21'E |  |
| *R. paradoxolophus* | Fenkeng Cave | Guilin City | Guangxi | 25°17'N | 110°21'E |  |
| *R. paradoxolophus* | Xipo Cave | Wuchuan County | Guizhou |  |  |  |
| *R. paradoxolophus* | Qingxuan Cave |  | Yunnan | 26°40'N | 100°12'E |  |
| *R. paradoxolophus* | Tianlong Cave | Dali County | Yunnan | 25°55'N | 100°05'E |  |
| *R. paradoxolophus* | Bianfu Cave | Yiliang County | Yunnan | 25°04'N | 103°23'E |  |
| *R. paradoxolophus* | Xiyou Cave | Fumin County | Yunnan | 25°09'N | 102°39'E |  |
| *R. paradoxolophus* | Xiaogou Cave | Yiliang County | Yunnan |  |  |  |
| *R. sinicus* | Yinhua Cave | Jiangle County | Fujian | 26°39'N | 117°34'E |  |
| *R. sinicus* | Bianfu Cave | Taining City | Fujian | 26°42'N | 117°30'E |  |
| *R. sinicus* | Longkong Cave | Yanshi County | Fujian | 25°12'N | 117°15'E |  |
| *R. sinicus* | Mine | Buyun Town, Shanghang County | Fujian | 25°15'N | 116°50'E |  |
| *R. sinicus* | Jinkuang Cave | Liancheng County | Fujian | 25°39'N | 116°53'E |  |
| *R. sinicus* | Liuhuang Cave | Liancheng County | Fujian | 25°52'N | 117°17'E |  |
| *R. sinicus* | Yuxi Cave | Mingxi County | Fujian | 26°21'N | 117°11'E |  |
| *R. sinicus* | Yinhua Cave | Jiangle County | Fujian | 26°39'N | 117°34'E |  |
| *R. sinicus* | Xing Cave | Qiliqiao Bridge, Wuyi Mountain | Fujian | 27°45'N | 117°30'E |  |
| *R. sinicus* | Mine, Chang ganzhou Road | Xincun Town, Wuyi City | Fujian | 27°37'N | 117°50'E |  |
| *R. sinicus* | Huanghuacong Cave | Yanzijiao Town, Wuyi Mountain | Fujian | 27°49'N | 117°43'E |  |
| *R. sinicus* | Penglai Cave | Yunfu Mount, Bolo County | Guangdong |  |  |  |
| *R. sinicus* | Fangkong | Luofu Mountain, Huizhou City | Guangdong | 23°16'N | 114°04'E |  |
| *R. sinicus* | Ni Cave | Xinyi County | Guangdong | 22°20'N | 110°57'E |  |
| *R. sinicus* | Shuili Cave | Xinyi County | Guangdong | 22°20'N | 110°58'E |  |
| *R. sinicus* | Kiln | Xinyi County | Guangdong | 22°15'N | 110°60'E |  |
| *R. sinicus* | Closed Cave | Ziyuan County | Guangxi |  |  |  |
| *R. sinicus* | Fenkeng Cave | Guilin City | Guangxi | 25°17'N | 110°21'E |  |
| *R. sinicus* | Fenkeng Cave | Guilin City | Guangxi | 25°17'N | 110°21'E |  |
| *R. sinicus* | Shui Cave | Guilin City | Guangxi |  |  |  |
| *R. sinicus* | Jiang Cave | Jiangkou County | Guizhou | 23°25'N | 110°15'E |  |
| *R. sinicus* | Xiashui Cave | Qiongzong County | Hainan | 19°04'N |  |  |
| *R. sinicus* | Hela Village | Hongmao Town, Qiongzong County | Hainan |  |  |  |
| *R. sinicus* | Diya Village | Five Finger Mountain | Hainan |  |  |  |
| *R. sinicus* | Fangkong Cave | Jianfeng Mountain | Hainan | 18°54'N | 109°42'E |  |
| *R. sinicus* | Cave | Yichang City | Hubei |  |  |  |
| *R. sinicus* | Longhushan Park | Yingtian City | Jiangxi | 28°04'N | 116°58'E |  |
| *R. sinicus* | Wanfu Cave | Jingdezhen County | Jiangxi | 28°37'N | 116°28'E |  |
| *R. sinicus* | Fufeng Cave | Emeishan | Sichuan | 29°34'N | 103°25'E |  |
| *R. sinicus* | Jiulao Cave | Emeishan | Sichuan | 29°33'N | 109°47'E |  |
| *R. sinicus* | Bailong Cave | Mile County | Yunnan | 24°12'N | 103°14'E |  |
| *R. pusillus* | Shidu Caves | Fangshan District | Beijing | 39°25'N | 115°17'E |  |
| *R. pusillus* | Sanqing Cave | Wanglaopu, Fangshan District | Beijing | 39°45'N | 115°45'E |  |
| *R. pusillus* | Longkong Cave | Yanshi County | Fujian | 25°12'N | 117°15'E |  |
| *R. pusillus* | Yinhua Cave | Jiangle County | Fujian | 26°39'N | 117°34'E |  |
| *R. pusillus* | Jinkuang Cave | Liancheng County | Fujian | 25°39'N | 116°53'E |  |
| *R. pusillus* | Ganru Cave | Liancheng County | Fujian |  |  |  |
| *R. pusillus* | Niutongguan Cave | Liancheng County | Fujian | 25°33'N | 116°59'E |  |
| *R. pusillus* | Chuqi Cave | Liancheng County | Fujian | 25°33'N | 116°60'E |  |
| *R. pusillus* | Liu Huang Cave | Liancheng County | Fujian | 25°52'N | 117°17'E |  |
| *R. pusillus* | Qixian Cave | Sha County | Fujian | 26°25'N | 117°39'E |  |
| *R. pusillus* | Yinhua Cave | Jiangle County | Fujian | 26°39'N | 117°34'E |  |
| *R. pusillus* | Jiuba Cave | Jiangle County | Fujian | 26°43'N | 117°29'E |  |
| *R. pusillus* | Cave of the Buddha | Dingushan Forest Eco system, | Guangdong |  |  |  |
| *R. pusillus* | Butterfly Cave | Yunfu Mount | Guangdong |  |  |  |
| *R. pusillus* | Fangkong | Luofu Mountain, Huizhou City | Guangdong | 23°16'N | 114°04'E |  |
| *R. pusillus* | Dianxian Cave | Xinyi County | Guangdong | 22°20'N | 110°57'E |  |
| *R. pusillus* | Shuili Cave | Xinyi County | Guangdong | 22°20'N | 110°58'E |  |
| *R. pusillus* | Rock crevice | Xinyi County | Guangdong | 22°15'N | 111°00'E |  |
| *R. pusillus* | Tongtian Cave | Xinyi County | Guangdong | 22°23'N | 110°72'E |  |
| *R. pusillus* | Chang Cave | Xinyi County | Guangdong | 22°17'N | 111°02'E |  |
| *R. pusillus* | Jishui Cave | Guilin City | Guangxi | 25°17'N | 110°21'E |  |
| *R. pusillus* | White Dragon Cave | Guilin City | Guangxi |  |  |  |
| *R. pusillus* | Seven Star Cave | Guilin City | Guangxi | 25°17'N | 110°18'E |  |
| *R. pusillus* | Closed Cave | Ziyuan County | Guangxi |  |  |  |
| *R. pusillus* |  | Liangjiang Town, Wuming County | Guangxi |  |  |  |
| *R. pusillus* | Shui Cave | Guilin City | Guangxi |  |  |  |
| *R. pusillus* | Shui Cave | Guilin City | Guangxi |  |  |  |
| *R. pusillus* | Jishui Cave | Guilin City | Guangxi | 25°17'N | 110°21'E |  |
| *R. pusillus* | Seven Star Cave | Guilin City | Guangxi | 25°17'N | 110°18'E |  |
| *R. pusillus* | Wulong Cave | Mashan County | Guangxi |  |  |  |
| *R. pusillus* | Shanjia Cave | Anlong County | Guizhou | 25°19'N | 105°05'E |  |
| *R. pusillus* |  | Leigong Mountain | Guizhou |  |  |  |
| *R. pusillus* | Fangkong Cave | Jianfeng Mountain | Hainan | 18°54'N | 109°42'E |  |
| *R. pusillus* |  | Yichang City | Hubei | 31°09'N | 111°08'E |  |
| *R. pusillus* | Shenlong Cave | Yichang | Hubei | 31°21'N | 110°30'E |  |
| *R. pusillus* | Yeren Cave | Shennong gja | Hubei | 31°55'N | 110°44'E |  |
| *R. pusillus* | Sihe Village | Langhe Town, Wudang Mountain | Henan |  |  |  |
| *R. pusillus* | Xianren Cave | Wuhan City | Hubei | 25°53'N | 114°30'E |  |
| *R. pusillus* | Longhushan Park | Yingtian City | Jiangxi | 28°04'N | 116°58'E |  |
| *R. pusillus* | Wanfu Cave | Jing dezhen County | Jiangxi | 28°37'N | 116°28'E |  |
| *R. pusillus* | Xiaya Cave | Yiyuan County | Shandong |  |  |  |
| *R. pusillus* | Mine | Buyun Town, Shanghang County | Shandong | 25°15'N | 116°50'E |  |
| *R. pusillus* |  | Yilai Mountain | Sichuan | 30°29'N | 103°24'E |  |
| *R. pusillus* |  | Luding County | Sichuan | 29°48'N | 102°14'E |  |
| *R. pusillus* | Qilai County | Qilai County | Sichuan |  |  |  |
| *R. pusillus* | Ahei Cave | Xishuangbanna | Yunnan |  |  |  |
| *R. pusillus* | Heshang Cave | Kunming, Fumin County | Yunnan | 25°12'N | 102°28'E |  |
| *R. pusillus* | Xiangshui Village | Fumin County | Yunnan |  |  |  |
| *R. pusillus* | Wangzhangya Cave | Dawei County | Yunnan |  |  |  |
| *R. pusillus* | Fengjing Cave | Jiuxiang County | Yunnan |  |  |  |
| *R. pusillus* | Jiuxiang Cave | Yiliang County | Yunnan |  |  |  |
| *R. pusillus* | Bailong Cave | Mile County | Yunnan | 24°12'N | 103°14'E |  |
| *R. pusillus* | Shaft Cave, near Bailong Cave | Mile County | Yunnan | 24°11'N | 103°21'E |  |
| *R. stheno* | Ahei Cave | Xishuang banna | Yunnan |  |  |  |
| *R. stheno* | Jinuo Cave | Xishuang banna | Yunnan | 21°58'N | 100°49'E |  |

---

**Table S2.** Mean land cover and land use attributes (and 20% and 80% percentiles) in the surroundings (<30km distance) of bat location and randomly selected points (see methods) within and outside China.

|  | **Bat Locations** |  |  | **Random in China** |  |  | **Random out China** |  |  |
| --- | --- | --- | --- | --- | --- | --- | --- | --- | --- |
|  | *Mean* | *20%* | *80%* | *Mean* | *20%* | *80%* | *Mean* | *20%* | *80%* |
| **Livestock Density (heads/km<sup>2</sup>)** |  |  |  |  |  |  |  |  |  |
| - Chickens | 781 | 183 | 931 | 703 | 160 | 841 | 317 | 15 | 317 |
| - Ducks | 256 | 18 | 294 | 202 | 10 | 264 | 26 | 0 | 16 |
| - Pigs | 99 | 42 | 163 | 107 | 43 | 148 | 18 | 0 | 21 |
| - Goats | 32 | 0 | 47 | 30 | 4 | 42 | 8 | 0 | 9 |
| - Sheep | 5 | 0 | 0 | 3 | 0 | 1 | 14 | 0 | 21 |
| - Cattle | 22 | 5 | 31 | 22 | 6 | 32 | 16 | 2 | 21 |
| **Forest Cover (%)** | 45 | 19 | 71 | 35 | 13 | 55 | 52 | 20 | 80 |
| **Forest Fragmentation (°)** | 73 | 54 | 96 | 83 | 68 | 98 | 61 | 35 | 90 |
| **Cropland (°)** | 20 | 5 | 36 | 24 | 6 | 38 | 30 | 3 | 59 |
| **People (cap/km<sup>2</sup>)** | 282 | 63 | 318 | 199 | 59 | 270 | 107 | 12 | 114 |
| **Villages/Human Settlements (°)** | 11 | 3 | 18 | 10 | 2 | 16 | 4 | 0 | 5 |

**Table S3.**
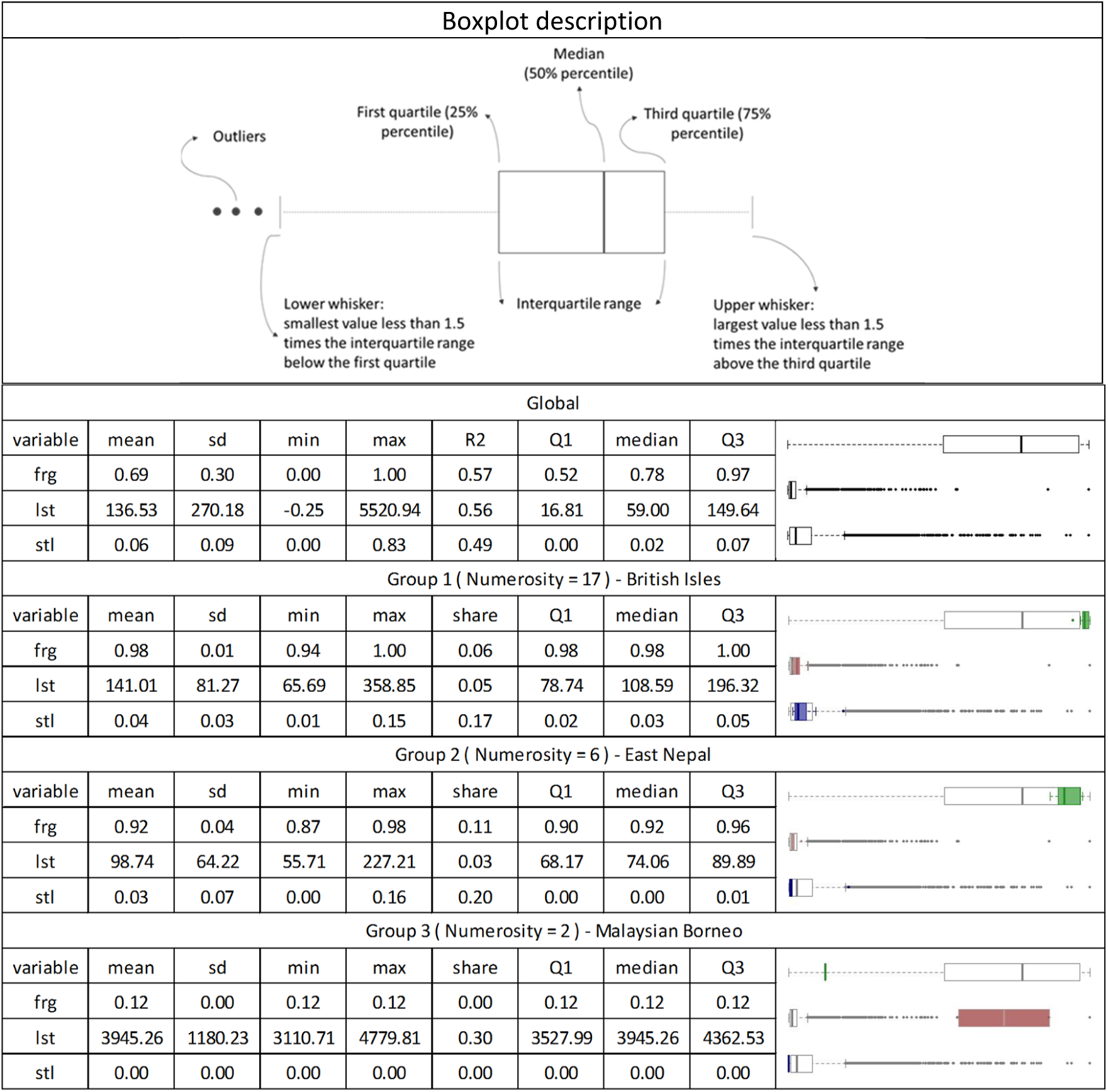
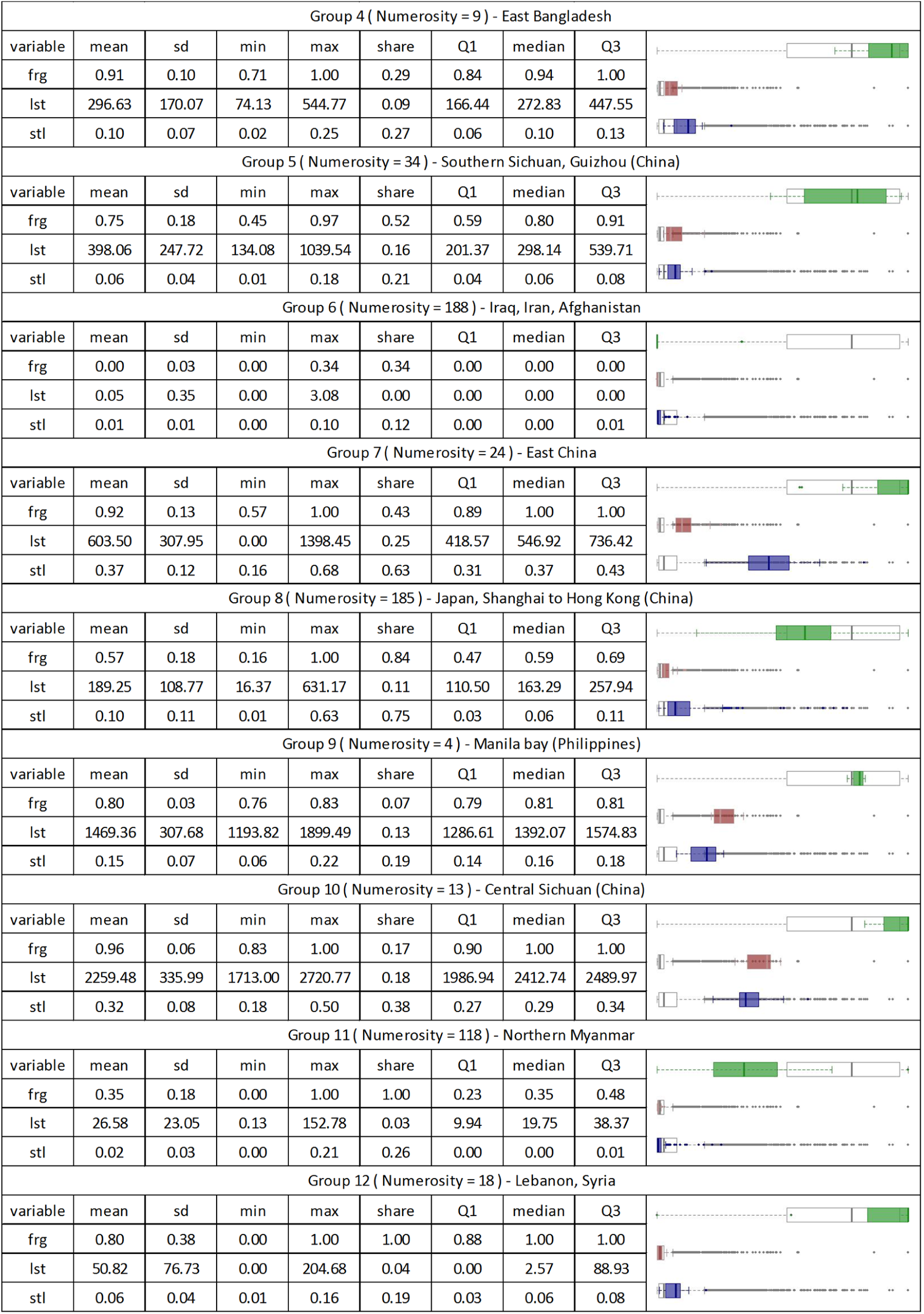
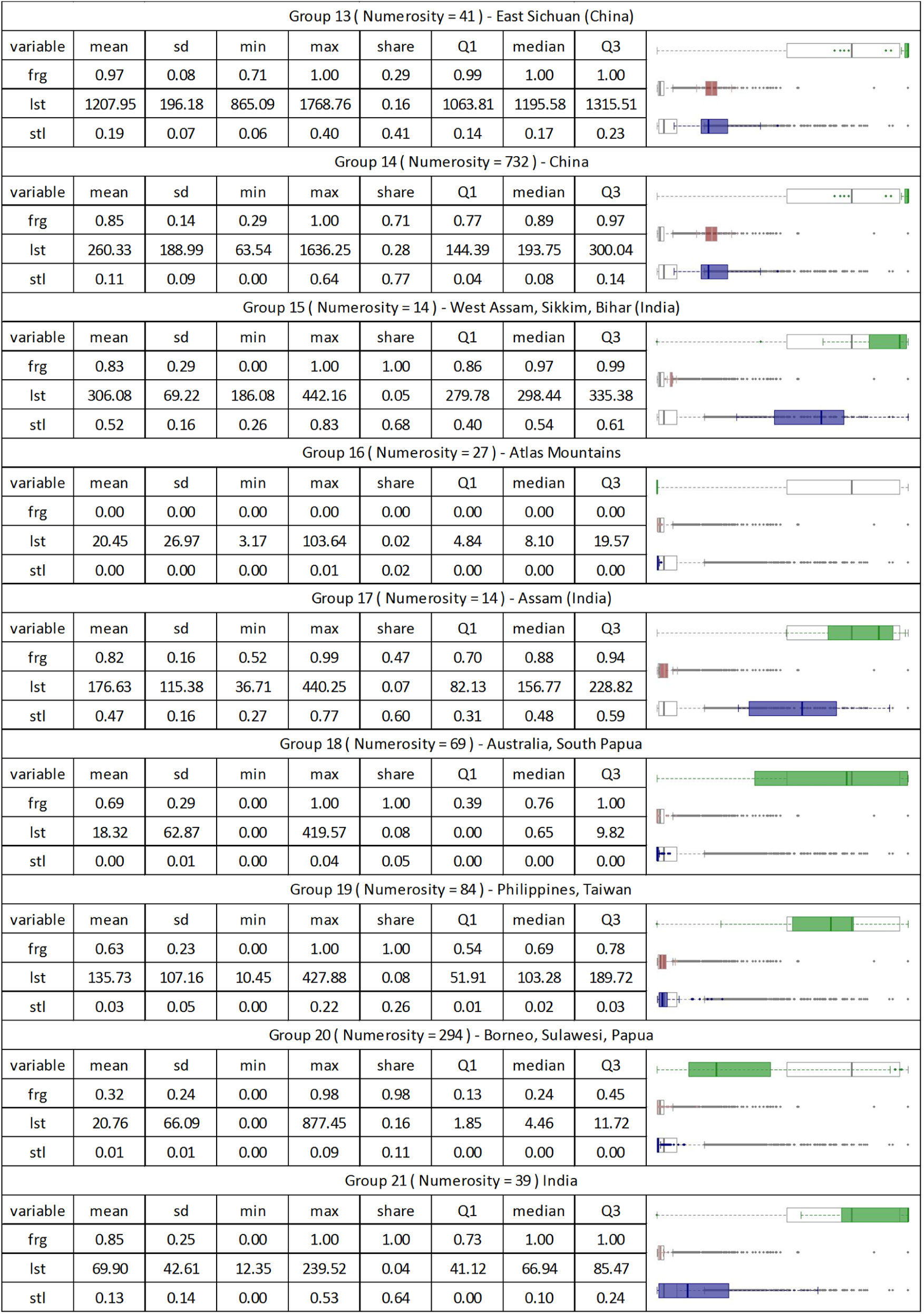
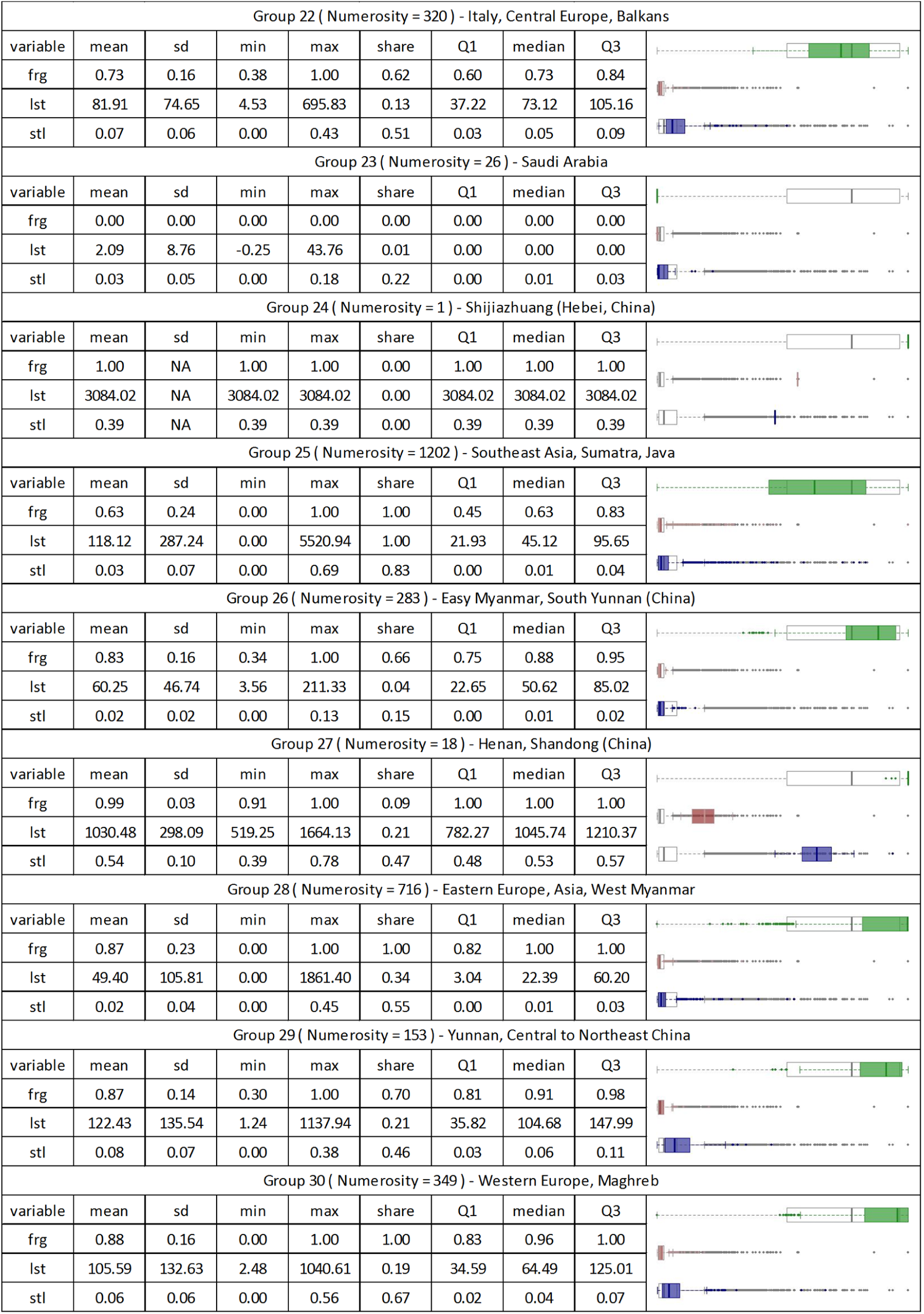
Report for the spatially constrained multivariate grouping analysis, showing group numerosity, descriptive statistics and boxplots, in comparison with the global benchmark. The share is calculated as the ratio of the variable range (min-max) which is occupied by the same variable in a group. Q1 and Q3 are respectively the first and third quartile, and sd is the standard deviation. The R2 parameter measures the indicator’s performance in grouping the sample, as relative reduction of the intra-group variability w.r.t. the global sample variability. Variables include fragmentation (frg), livestock (lst), and settlements (stl). Boxplots are an intuitive tool to visually recap statistical distributions. The meaning of the elements of boxplots is described in the figure below. In the report, the first and third quartile are expressed respectively as Q1 and Q3. The high number of outliers for livestock and settlements is typical for high-skewness distributions. The boxplots show well how the indicator distributions shift and narrow from the global dataset to single groups. Although in some the standard deviation of one or more indicators is higher than the global benchmark, each group can be considered as homogeneous for at least one parameter.

**Figure S1.**
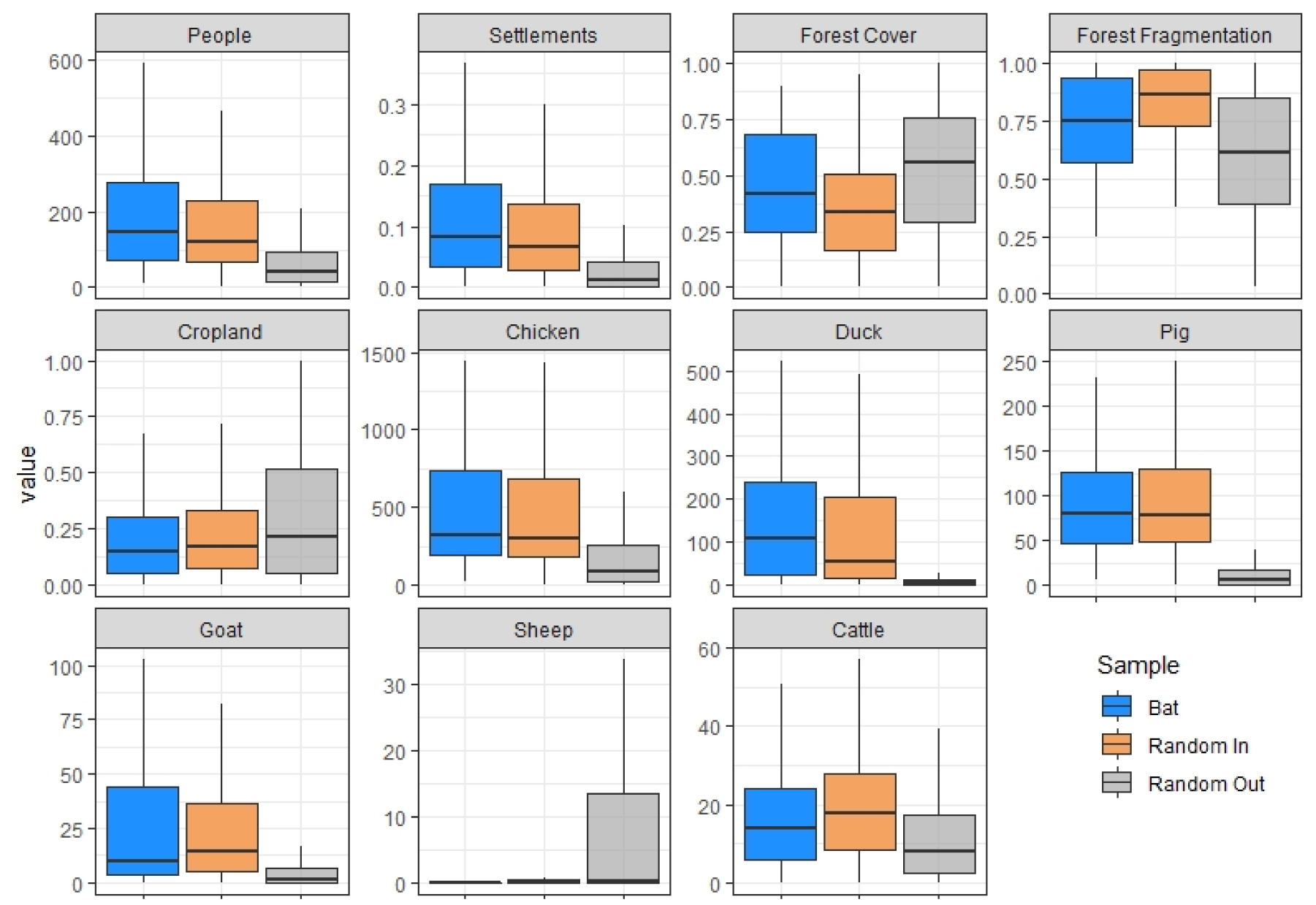
Boxplots of the distribution data of indicators. In blue are reported data in bat locations, in orange data in China, in grey data outside China.

**Figure S2.**
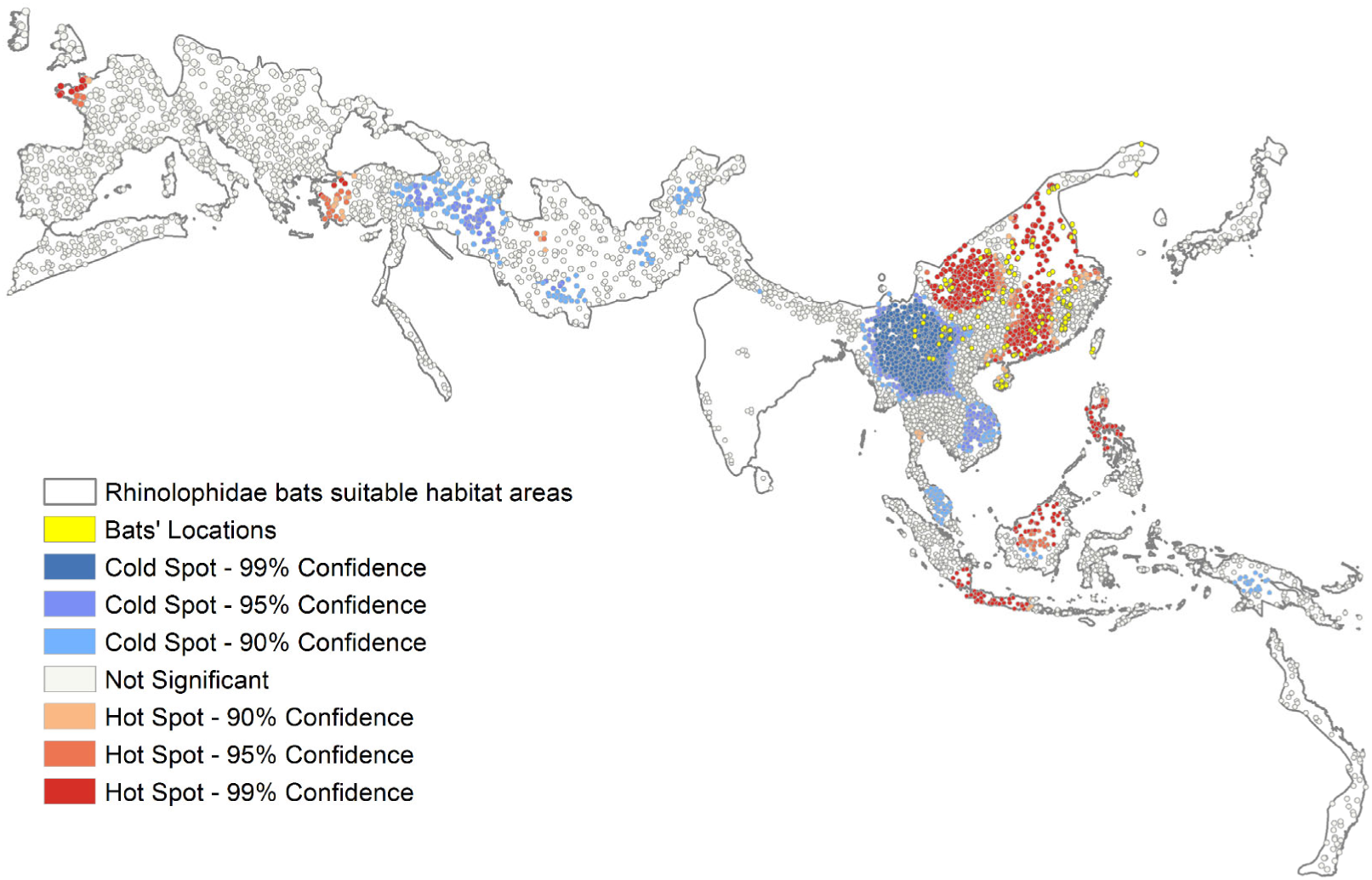
Hotspots and coldspots of chicken density.

**Figure S3.**
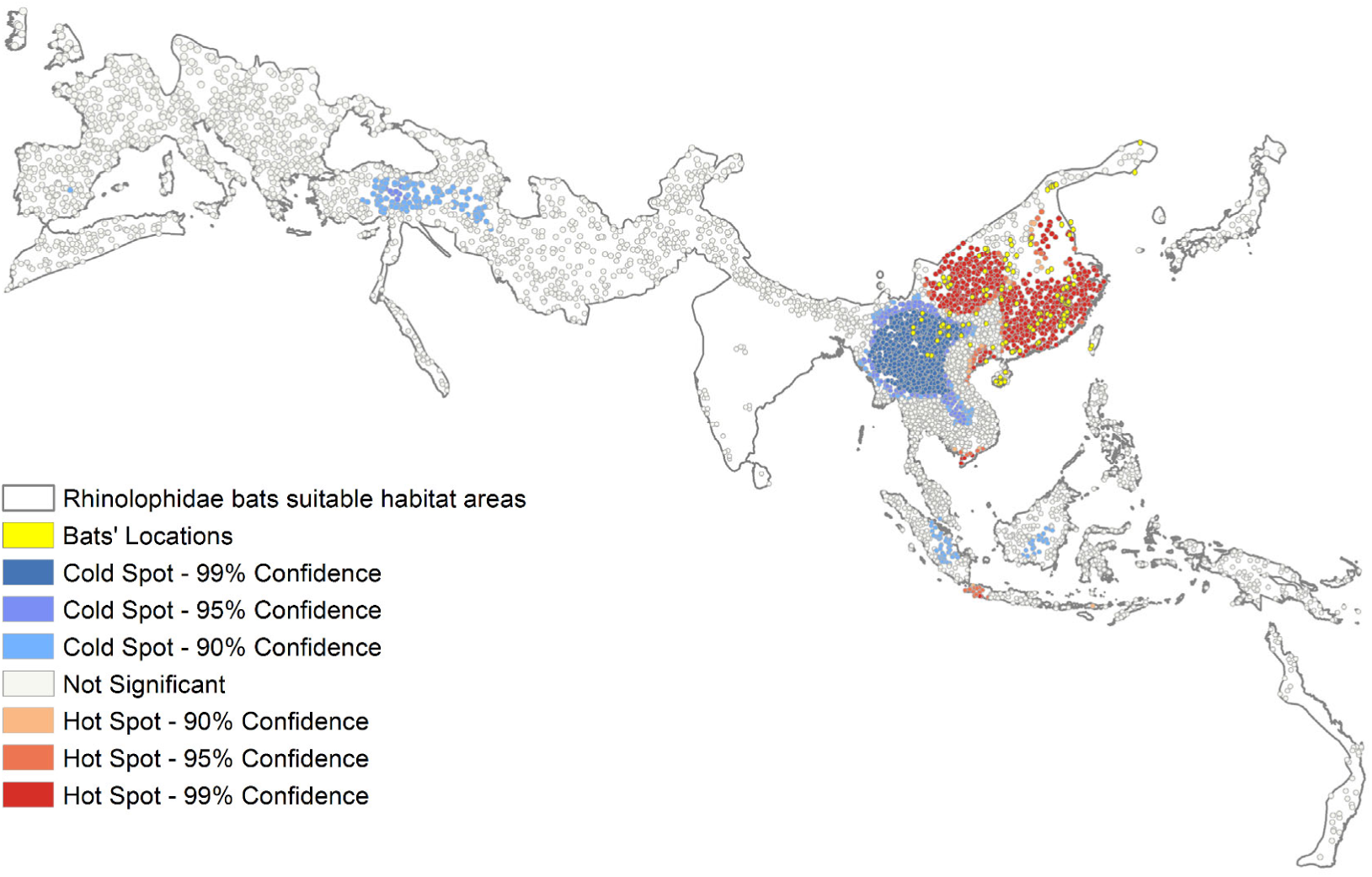
Hotspots and coldspots of duck density.

**Figure S4.**
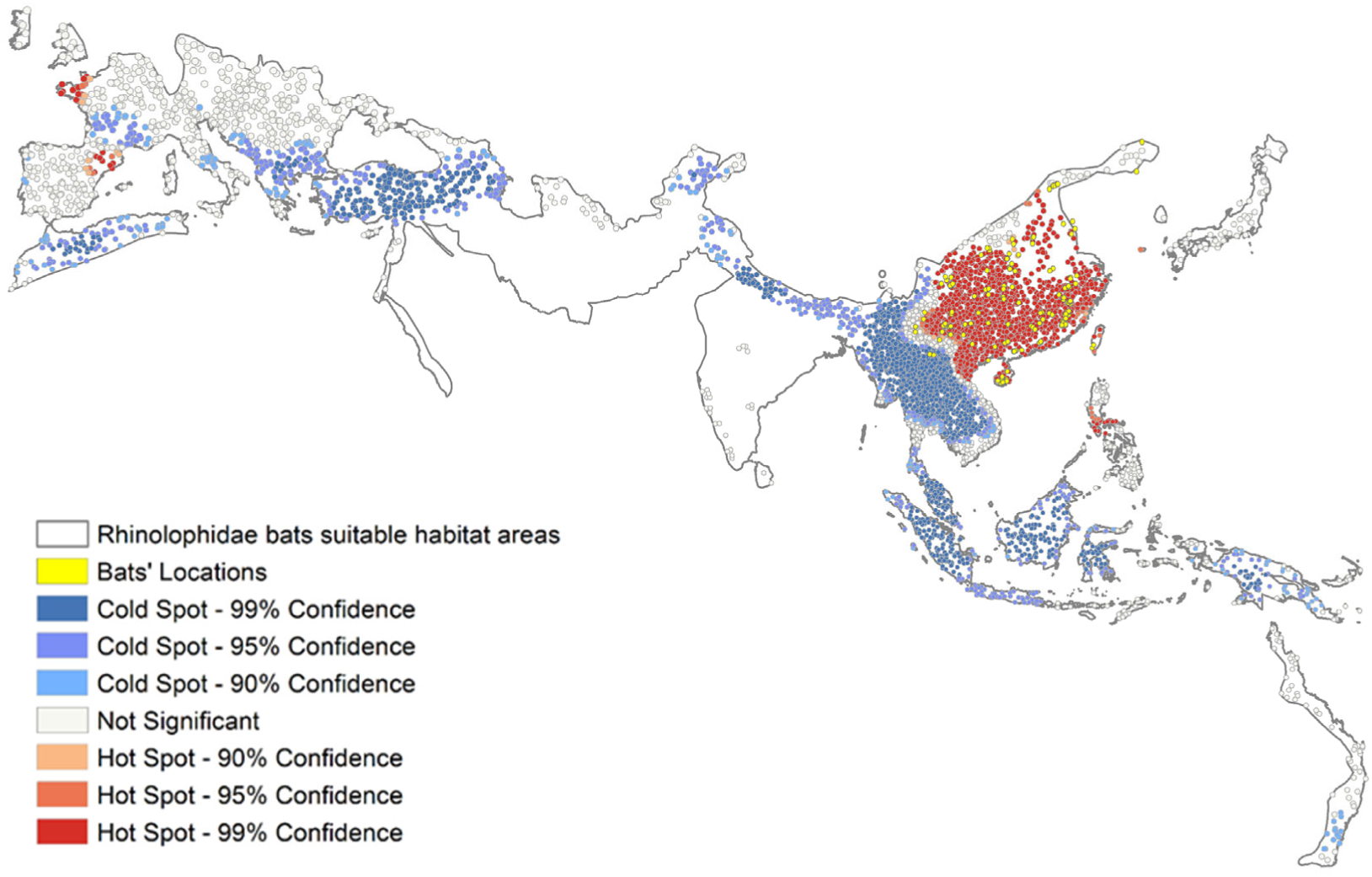
Hotspots and coldspots of pig density.

**Figure S5.**
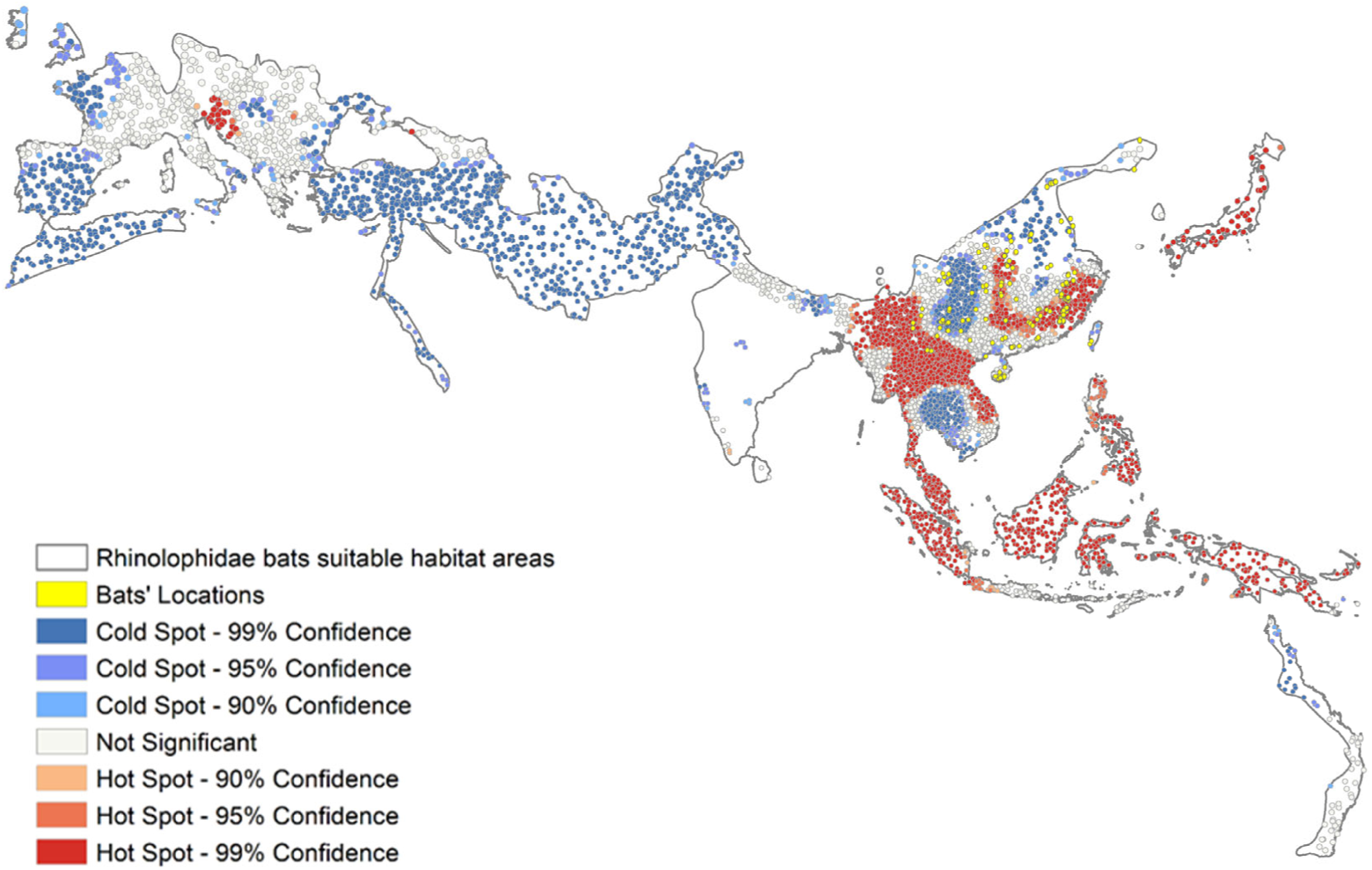
Hotspots and coldspots of forest cover

**Figure S6.**
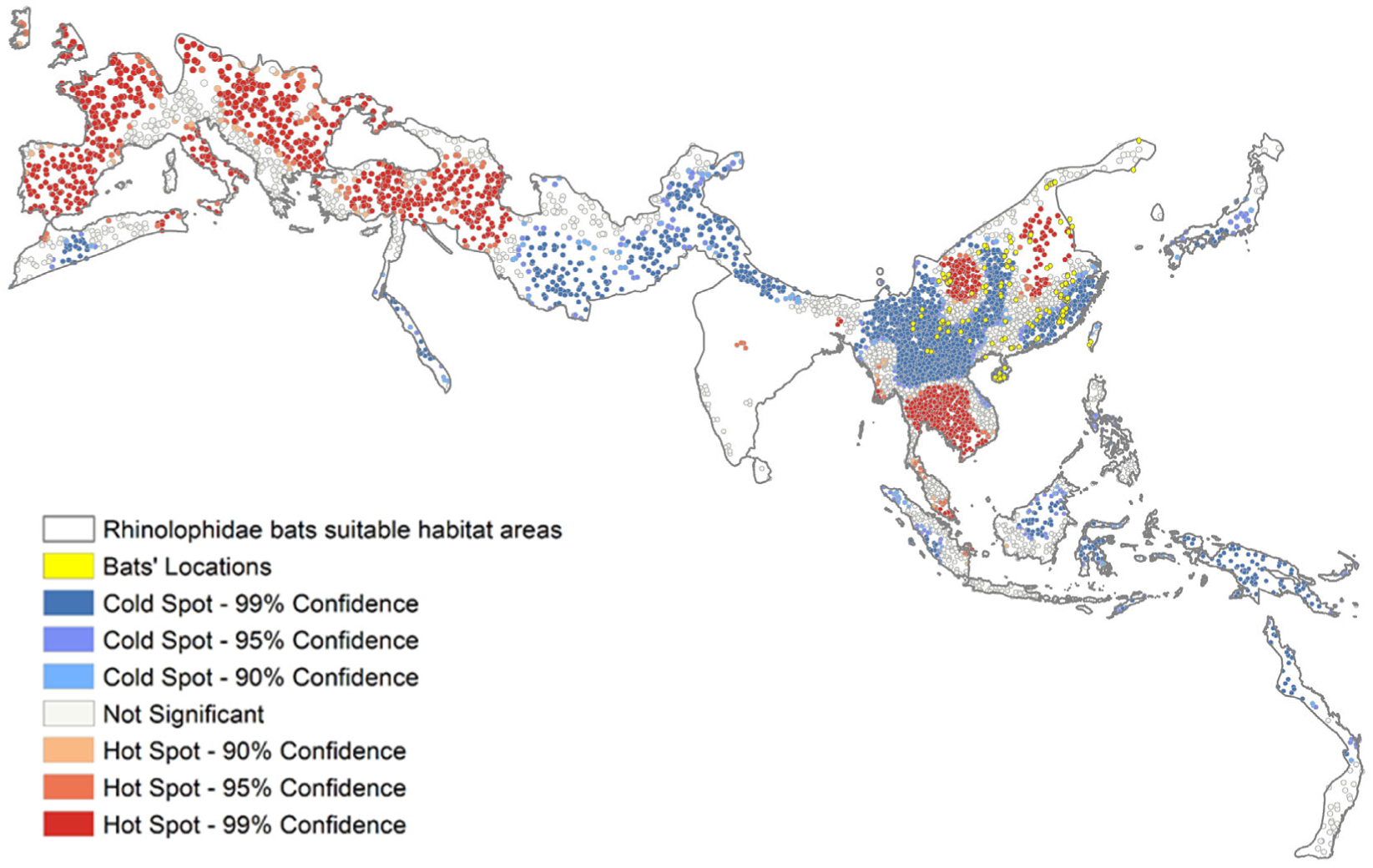
Hotspots and coldspots of cropland cover.

**Figure S7.**
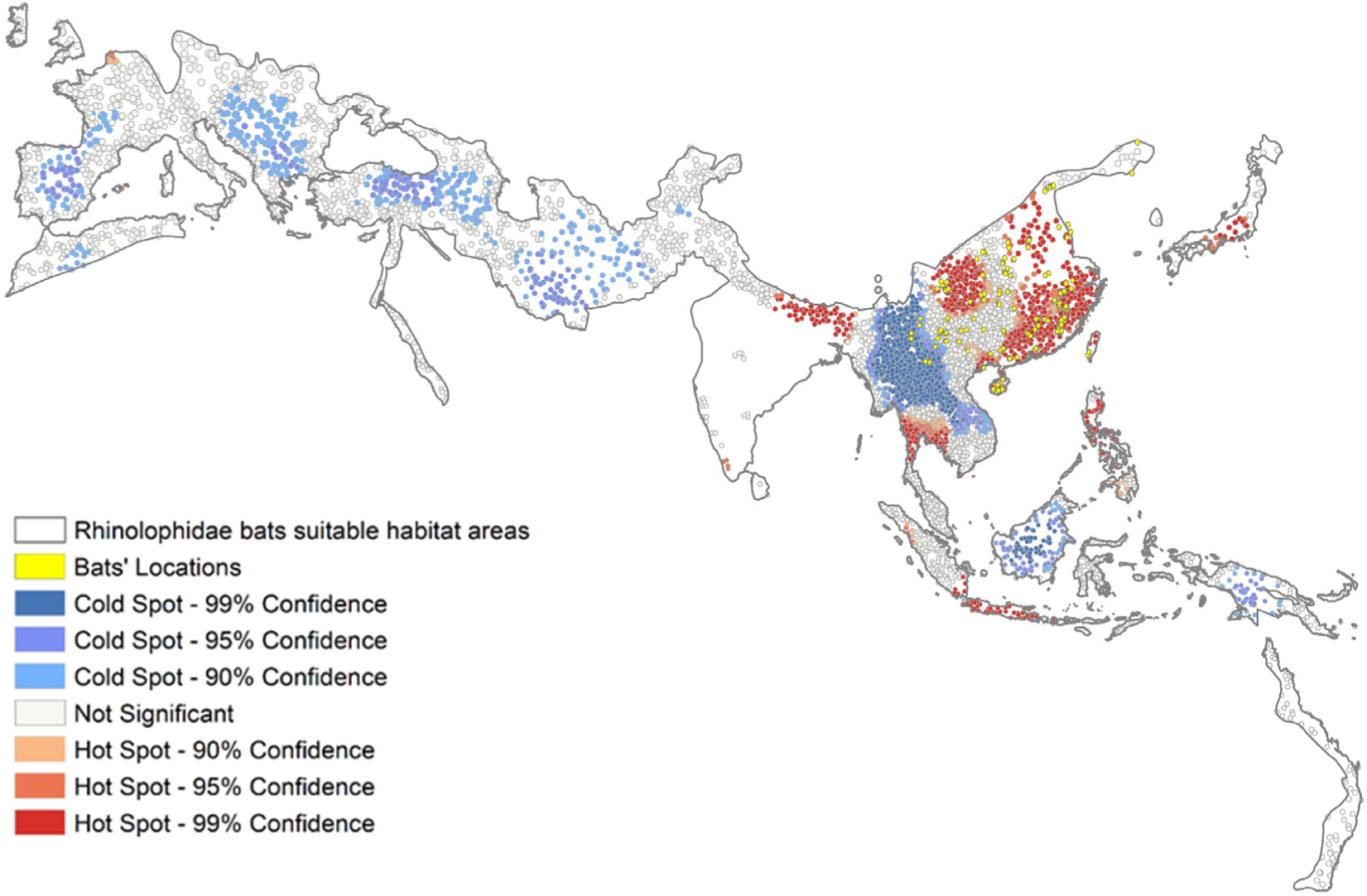
Hotspots and coldspots of population density.

**Figure S8.**
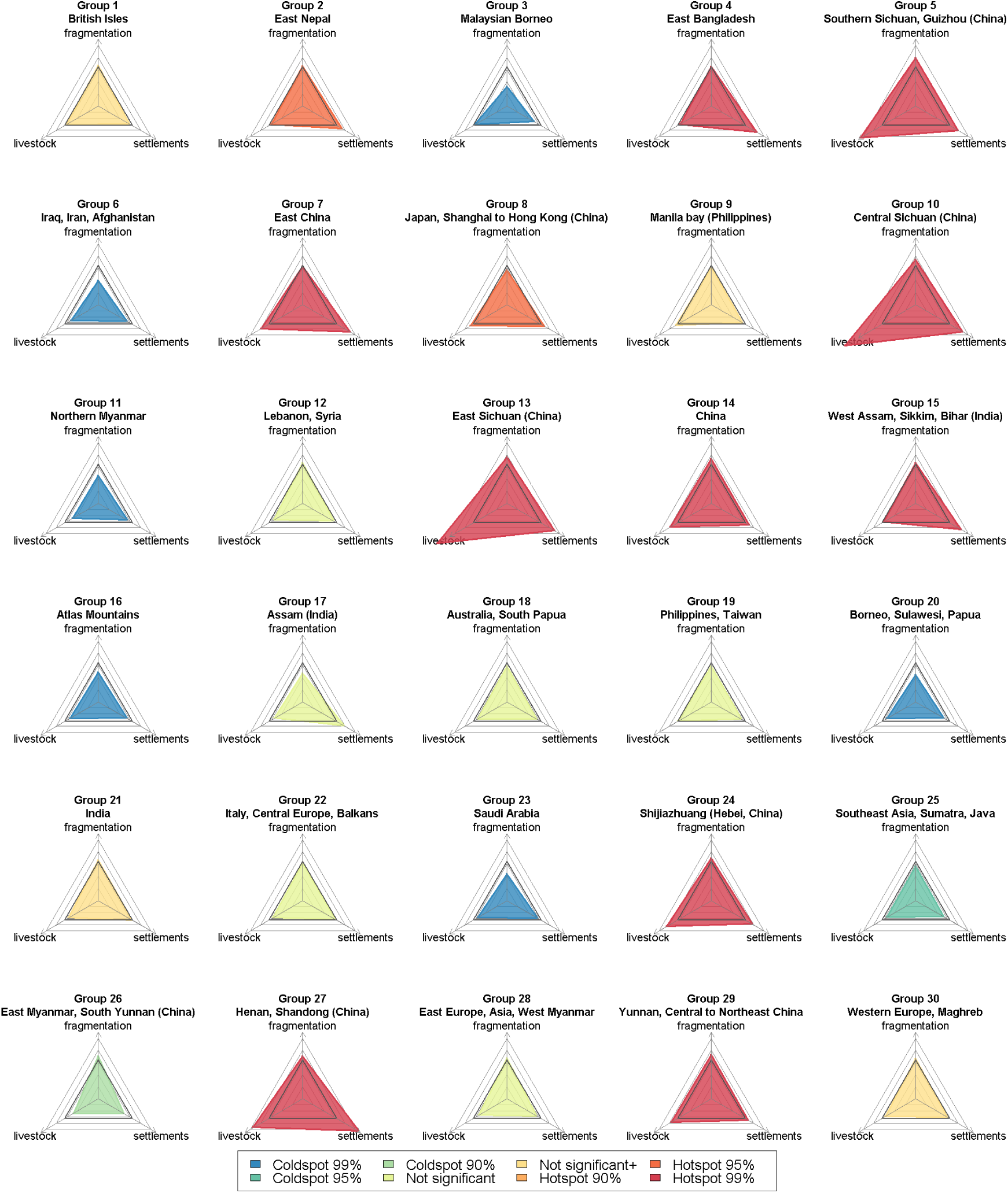
Group analysis. Trajectories of risk.

## Notes

### Competing Interest Statement

The authors have declared no competing interest.

### Funding Statement

M.C.R and N.G. are supported by Cariplo Foundation (SusFeed project 0737 CUPD49H170000300007). D.T.S.H. is supported by Rutherford Discovery Fellowship RDF MAU1701.

### Author Declarations

This has no patient data.

